# Phenotypic Selectivity of Artificial Intelligence-enhanced Electrocardiography in Cardiovascular Diagnosis and Risk Prediction

**DOI:** 10.1101/2025.07.01.25330665

**Authors:** Philip M. Croon, Lovedeep S. Dhingra, Dhruva Biswas, Evangelos K Oikonomou, Rohan Khera

## Abstract

**Introduction:** Artificial intelligence (AI)-enhanced electrocardiogram (ECG) models are designed to detect specific anatomical and functional cardiac abnormalities. Understanding the selectivity of their phenotypic associations is essential to inform their clinical use. Here, we sought to assess whether AI-ECG models function as condition-specific classifiers or broader cardiovascular risk markers.

**Methods:** We included four distinct study populations, drawn from both electronic health records (EHR) and prospective cohort studies. We deployed six image-based AI-ECG models, including five validated models for the detection of left ventricular systolic dysfunction (LVSD), aortic stenosis (AS), mitral regurgitation (MR), left ventricular hypertrophy (LVH), a composite model for structural heart disease (SHD), and a negative control AI-ECG model for biological sex. Additionally, we developed six experimental models designed to identify non-cardiovascular conditions. Diagnosis codes from EHR and cohorts were transformed into interpretable phenotypes using a phenome-wide association study (PheWAS) framework. We assessed associations of AI-ECG probabilities with cross-sectional phenotypes using logistic regression, and with new-onset cardiovascular diseases using Cox regression. Pearson correlation coefficients were calculated to compare phenotypic signatures.

**Results:** The study included one random ECG from 233,689 individuals (mean age 59±18 years, 130,084 [56%] women) across sites. Each of the five AI-ECG models was more likely to be associated with cardiovascular phenotypes compared with other phenotype groups (odds ratios ranging from 2.16 to 4.41, p<10□), while the sex model did not show a similar pattern. All AI-ECG models were significantly associated with their respective target phenotype, but also showed similar or stronger associations with a broad range of other cardiovascular phenotypes. Phenotypic associations were similar across AI-ECG models trained for different conditions, which was not observed in models for non-cardiovascular conditions. Correlation of phenotype association patterns between models was high (r = 0.65–0.99). This pattern was consistent across all models, external datasets, and in both cross-sectional and prospective analyses.

**Conclusions:** Despite being developed to detect specific cardiovascular conditions, AI-ECG models detect the presence and predict the future development of a broad range of cardiovascular diseases with similar propensity. This challenges their role as binary diagnostic tools and instead supports their use as broader cardiovascular biomarkers.

## INTRODUCTION

Artificial intelligence (AI) has expanded the utility of the electrocardiogram (ECG), enabling the detection of subtle disease signatures previously considered unrecognizable even by experts.^1,2^ Several AI-ECG models have now been extensively validated for the detection of various specific functional and structural cardiovascular conditions, including left ventricular systolic dysfunction (LVSD), valvular heart disease (VHD), and left ventricular hypertrophy (LVH).^3–7^ Moreover, AI-ECG has demonstrated effectiveness as a digital biomarker capable of predicting new-onset cardiovascular diseases (CVD).^8–11^ These developments have led to the FDA approval for AI-ECG algorithms designed to detect LVSD and hypertrophic cardiomyopathy, marking its transition from research into clinical application.^12^

While AI-ECG models have been developed and evaluated as condition-specific tools, an assessment of their selectivity in phenotypic associations is required to guide their clinical use. Many CVDs exhibit shared pathophysiological mechanisms and risk factors, which may result in similar conduction and rhythm alterations.^10,13^ Visible ECG abnormalities, such as ST-segment changes or prolonged QT intervals, are also frequently associated with multiple cardiac diseases, reflecting overlapping pathological processes.^14–16^ As a result, AI-ECG models may not differentiate between distinct disease entities, as suggested by a series of models designed for individual conditions, but instead identify ECG signatures representing broader cardiovascular pathology. With the ongoing transition of AI-ECG into clinical practice, these potential limitations must be systematically evaluated to clarify their clinical implications and ensure safe, effective deployment.^17^

In this study, we evaluated whether image-based AI-ECG models trained on specific cardiovascular disease labels exhibit selective phenotypic association patterns for cross-sectional and new-onset cardiovascular disorders across four diverse cohorts, including a large tertiary hospital, a group of four community hospitals, an outpatient medical network, and the prospective UK Biobank. We also included a classification model for biological sex and seven experimental models as negative control outcomes for cardiac phenotypic patterns potentially detectable on AI-ECG.

## METHODS

The Yale Institutional Review Board approved the study protocol and waived the requirement for informed consent, as the study involves secondary analysis of pre-existing data. The data underlying this study cannot be shared publicly due to patient privacy concerns and institutional regulations.

### Evaluation of AI-ECG Models for Cardiovascular Disease

We leveraged a range of AI-ECG models that were previously developed and extensively validated in multinational cohorts to detect distinct cardiovascular abnormalities. These cardiovascular disease models were trained and validated for specific labels including: (1) LVSD, defined as a left ventricular ejection fraction (LVEF) < 40% on echocardiography, left-sided valvular heart diseases, namely (2) AS and (3) MR, graded as mild to severe based on echocardiographic guidelines, (4) severe LVH, defined by an interventricular septal diameter at end-diastole > 15 mm, combined with moderate to severe (grade II/III) left ventricular diastolic dysfunction, and (5) composite SHD including LVSD, valvular heart disease and LVH.^18^

The development and external validation of these models, including cohort characteristics, has been described in prior studies.^3,19^ All models were developed in the same group of individuals who had undergone an electrocardiogram and echocardiogram concurrently (within 30 days of each other) in the Yale New Haven Hospital (YNHH).

The individual disease label models are all convolutional neural networks (CNN) based on the EfficientNet-B3 architecture, which consists of 384 layers and over 10 million trainable parameters. The model input consists of 300×300 pixel image representations of standard 12-lead ECGs. To enhance label efficiency in model development, CNNs were initialized with weights from a self-supervised, contrastive learning framework designed to recognize patient-specific ECG patterns independent of clinical interpretation.^20^ Fine-tuning was performed in two stages. In the initial stage, only the final two layers of the network were unfrozen and optimized, followed by a second stage in which all layers were unfrozen to allow full model adaptation to the task-specific objective. This staged training approach was used to improve optimization stability and efficiency, while preserving the model’s capacity to update all weights based on the disease-specific signal. The AI-ECG model for detecting composite structural heart disease, called PRESENT-SHD, employs an XGBoost classifier that integrates predictions from individual CNNs trained to detect LVSD, valvular heart disease, and LVH, along with an individual’s age and sex as input features.^19,21^ All models were externally validated across diverse clinical and community-based settings and demonstrated state-of-the-art performance.

### Development and Evaluation of Experimental AI-ECG Models

To evaluate whether the signature of an AI-ECG model trained on a non-cardiovascular, biological label differs from that of one trained to detect cardiovascular diseases, we developed an AI-ECG model to define biological sex from the ECG, using the same methodology and cohort as the disease-specific models described in the previous paragraph. Such models have previously been proposed, and this model demonstrated excellent performance (area under the receiver operating curve (AUROC)>0.90) across all cohorts, including internal validation in the YNHH cohort and external validation in community hospitals, outpatient clinics, and the UK Biobank **(Supplementary Figure 1, Supplementary Table 1)**.^22^

To examine whether the signatures of the AI-ECG models are dependent on the training label rather than consistently suggesting cardiovascular pathology, we developed multiple experimental models to serve as negative controls for the cardiovascular association of AI-ECG models. We used the same methodology and development population as the validated AI-ECG models for cardiovascular disease detection. First, we developed a model to classify whether an ECG was recorded during an odd-numbered month. Next, we derived 6 experimental models designed to detect the occurrence of non-cardiovascular disease labels, including viral respiratory infection, transport accident, headache, fracture of the lower leg, being bitten by a dog, and dermatophytosis. These conditions were defined based on the presence of diagnosis codes up to one year before or after the ECG **(Supplementary Table 2)**. None of these experimental models outperformed random chance in the development cohort (AUROC ∼0.50), confirming their role as negative control outcomes for AI-ECG **(Supplementary Table 3, Supplementary Figure 2)**.

### Data Sources

We included individuals from multiple cohorts. The primary dataset was sourced from YNHH, a large tertiary medical center in Connecticut, USA. For external validation, we included: (i) four community-based hospitals in the Yale New Haven Health System (YNHHS): Bridgeport, Greenwich, Lawrence + Memorial and Westerly Hospitals, (ii) Northeast Medical Group spanning multiple outpatient clinics, and (iii) participants from the UK Biobank cardiovascular imaging substudy with an ECG recorded at enrollment. Extensive health and biometric data were available through linkage with National Health Service (NHS) electronic health records. ECGs included in the cohorts from the YNHH, community hospitals, and outpatient clinics were recorded as part of routine clinical care from 2013 to 2024, while the ECGs from UK Biobank participants were collected between 2014 and 2020. For all cohorts, diagnostic codes, classified as International Classification of Diseases (ICD)-10 codes, along with the first recorded dates, were extracted.

### Study population

In the cohort derived from the YNHH, individuals who were part of the model development were excluded from our analysis. For the cohorts that originated from the YNHH, community hospitals, and outpatient clinics, we selected individuals with at least one ECG as part of routine clinical care. To ensure adequate follow-up for disease ascertainment, we applied a one-year blanking period starting from each patient’s first hospital encounter. We then randomly selected one ECG recorded after this period per individual, allowing for consistent follow-up and representation across the spectrum of disease stages. For the UK Biobank, we included all participants with an available ECG, using the earliest recorded ECG per individual. To define the baseline characteristics of the cohorts, we used patient demographics and diagnostic codes **(Supplementary Table 4).**

### Exploring Model Associations with Clinical Phenotypes

We conducted a phenome-wide association study (PheWAS) to explore cross-sectional associations between AI-ECG model predictions and a wide range of clinical phenotypes derived from diagnostic codes. PheWAS is an approach used to systematically evaluate associations between a specific genetic or clinical trait and a wide range of phenotypes.^23^ It has previously been used to identify novel disease associations, uncover comorbid conditions, and explore the phenotypic spectrum associated with biomarkers or genetic variants.^24–26^

For cross-sectional analysis, only diagnostic codes recorded before or on the day the ECG was recorded were considered, while those recorded afterwards were excluded. Additionally, to maintain statistical robustness, only phenotypes ≥20 occurrences were included in the analysis. These diagnostic codes were transformed into interpretable clinical phenotypes (e.g. heart failure, aortic valve disease) and their corresponding phenotype group (e.g. cardiovascular, respiratory) using the publicly available phecode catalog.^27,28^ The use of phenotypes as representations of clinical diagnosis codes and the reproducibility of phenotype associations have been validated in several prior reports.^23,29–31^ Patients with multiple coexisting conditions were included in each relevant phenotype category, ensuring our cross-sectional associations reflect real-world multimorbidity. However, to evaluate how concomitant diseases influence our findings, we performed a sensitivity analysis in which we established the cross-sectional associations on cohorts successively restricted to individuals with no more than one, then no more than two, and finally no more than three of the four key target phenotypes.

We defined phenotypes as either on-target, reflecting the specific condition that most closely approximates the disease that each model was trained to detect, or off-target, representing any other disease the model associated with. The target phenotypes were defined in consensus with a panel of three clinical cardiologists, who reviewed the AI-ECG models and available phecodes to identify those most closely aligned with each phenotype. Target phenotype names for each specific heart disease model were defined as follows: (1) heart failure for the LVSD model, (2) aortic valve disease for the AS model, (3) mitral valve disease for the MR model, (4) cardiomegaly, the phecode-derived phenotype most closely aligned with LVH, subsequently referred to as LVH and (5) a composite outcome that encompasses all of the above phenotypes for the SHD model **(Supplementary Table 5)**. Although these labels are not exhaustive, they provide a reproducible and representative framework for assessing disease-specific patterns of association and are clinically meaningful targets that align with prior work by our group and others.^8,19,32^

To assess whether phenotypic associations with cardiovascular phenotypes were consistent across study cohorts, we compared pairwise associations of effect estimates between the YNHH and external validation sites. We visualized the strength and direction of phenotype associations using volcano plots. To compare model-specific associations with phenotypes, we generated a heatmap of ORs between each AI-ECG model and the five target cardiovascular phenotypes. Next, we visualized the correlation of their phenotype association profiles across the same five target phenotypes, in a second set of heatmaps.

To assess the phenotypic overlap of AI-ECG models trained to detect conditions with distinct pathophysiology but overlapping clinical manifestations, we conducted a sensitivity analysis comparing the LVH and AS models from the main study with a previously published and validated hypertrophic cardiomyopathy (HCM) model with conditions that represent their target conditions.^33^

### Association of AI-ECG with New-Onset Cardiovascular Disease

To evaluate the prognostic value of AI-ECG model predictions for new-onset cardiovascular disease, we conducted time-to-event analyses using AI-ECG model predictions, including those for cardiovascular disease and biological sex, as independent variables and the predefined target phenotypes as dependent variables. In addition to individual conditions, we included a composite endpoint comprising all 5 target phenotypes. This resulted in five outcomes for which separate Cox proportional hazards models were fitted: (1) heart failure, (2) aortic valve disease, (3) mitral valve disease, (4) LVH, and (5) a composite of these conditions. To ensure analyses were limited to an at-risk sample, individuals with any of these diagnoses recorded before or on the day of the ECG acquisition were excluded.

### Statistical Analysis

The characteristics of the study population, the exposures, and the outcomes were summarized using means and standard deviations (SD) for continuous variables and counts and percentages for categorical variables.

Cross-sectional associations between AI-ECG model predictions and clinical phenotypes were assessed using age and sex adjusted logistic regression, where each phenotype was the dependent variable and individual AI-ECG probabilities were the key independent variable. These were reported as odds ratios (ORs) and 95% confidence intervals (95% CI). These analyses were conducted using the PheWAS R package (version 0.99.6.1).^34,35^

To evaluate whether the associations within phenotype groups identified by AI-ECG models exceeded what would be expected by chance, we compared the observed proportion of significant associations of the included models with the expected proportions using Fisher’s Exact test, with corresponding ORs and p-values reported. The predicted model probabilities from all models were logit-transformed and standardized for all analyses. To assess the reproducibility of individual phenotype associations across healthcare settings, we computed Pearson correlation coefficients between ORs and compared the results across cohorts.

To evaluate similarity in phenotypic association patterns across AI-ECG models, we first calculated pairwise Pearson correlation coefficients between ORs across the five predefined target phenotypes, separately within each cohort. Next, we repeated this analysis including all cardiovascular phenotypes. The resulting correlation matrices quantified the pairwise concordance in OR distributions across models, reflecting overlap in their phenotypic association profiles.

In assessing new-onset diseases, we developed age and sex adjusted Cox proportional hazards models with the models’ predictions as independent variables and the time to the occurrence of the first event as the dependent variable. Follow-up time was defined from the date of the ECG to the earliest occurrence of one of the following events: (i) onset of the tested phenotype, (ii) death, (iii) loss to follow-up, or (vi) day of data extraction, with a maximum follow-up time of five years. For the Cox models, hazard ratios (HRs) and 95% confidence intervals (CIs) were reported. To confirm that proportional-hazards and linearity assumptions held across all models, outcomes, and cohorts, we (1) plotted Schoenfeld residuals over time and (2) fitted five-knot restricted cubic splines for each predictor. Visual inspection of the disease-specific models’ diagnostics revealed no meaningful violations **(Supplementary Figure 3)**.

To control for multiple comparisons, we applied Bonferroni correction, with an adjusted significance threshold based on the number of phenotypes analyzed in each cohort. All statistical analyses were conducted using R (version 4.2.1) and Python (version 3.12.5).

## RESULTS

### Study Population

There were 235,685 individuals with at least one ECG of antecedent care within the YNHH. Of these, 116,540 were eligible for analysis, based on none of their data had been used in the model development and validation of prior AI-ECG models. These individuals had a mean age of 55.5□±□19.1 years and 65,920 (56.6%) were female, 69,560 (59.7%) self-identified as non-Hispanic White, 22,912 (19.7%) as non-Hispanic Black, and 18,130 (15.6%) as Hispanic. Among these participants, hypertension was present in 55,828 (47.9%), diabetes in 20,902 (17.9%), and heart failure in 8,611 (7.4%) individuals based on the presence of health encounters with corresponding diagnoses for these conditions in the period preceding the index ECG **(Table 1)**.

**Table 1:** Baseline characteristics of the included cohorts. This table presents the baseline characteristics of all cohorts and prevalence of both cross-sectional and new-onset key phenotypes. Baseline characteristics are based on demographics and ICD codes (**Supplementary Table 4**). Phenotypes are based on the publicly available PheWAS catalog which uses ICD codes and phenotype mapping (**Supplementary Table 5**). Binary variables are expressed as n (%), while continuous variables are reported as mean (Standard deviation). Abbreviations: FU follow-up; iqr, inter-quartile range.

|  | **Yale New**<br>**Haven Hospital** | **Community**<br>**Hospitals** | **Outpatient**<br>**clinics** | **UK**<br>**Biobank** |
| --- | --- | --- | --- | --- |
| **Total N** | 116540 | 63790 | 11005 | 42354 |
| Age | 55.5 ± 19.1 | 61.1 ± 19.5 | 61.4 ± 15.0 | 64.1 ± 7.8 |
| Sex Female | 65920 (56.6%) | 36088<br>(56.6%) | 6241<br>(56.7%) | 21835<br>(51.6%) |
| Race/Ethnicity |  |  |  |  |
| White | 69560 (59.7%) | 41401<br>(64.9%) | 8696<br>(79.0%) | 40893<br>(96.6%) |
| Black | 22912 (19.7%) | 7760 (12.2%) | 890 (8.1%) | 305<br>(0.7%) |
| Hispanic | 18130 (15.6%) | 11687<br>(18.3%) | 928 (8.4%) |  |
| Missing | 2624 (2.3%) | 1325 (2.1%) | 268 (2.4%) | 116<br>(0.3%) |
| Asian | 2054 (1.8%) | 958 (1.5%) | 146 (1.3%) | 603<br>(1.4%) |
| Others | 1260 (1.1%) | 659 (1.0%) | 77 (0.7%) | 437<br>(1.0%) |
| Heart Failure n (%) | 8611 (7.4%) | 5506 (8.6%) | 408 (3.7%) | 225<br>(0.5%) |
| Acute Myocardial<br>Infarction n (%) | 1617 (1.4%) | 1351 (2.1%) | 161 (1.5%) | 120<br>(0.3%) |
| Ischemic Heart Disease n<br>(%) | 9703 (8.3%) | 6434 (10.1%) | 1013 (9.2%) | 2233<br>(5.3%) |
| Stroke n (%) | 7710 (6.6%) | 5636 (8.8%) | 593 (5.4%) | 488<br>(1.2%) |
| Hypertension n (%) | 55828 (47.9%) | 32021<br>(50.2%) | 6800<br>(61.8%) | 6269<br>(14.8%) |
| Diabetes n (%) | 20902 (17.9%) | 12324<br>(19.3%) | 1793<br>(16.3%) | 1340<br>(3.2%) |
| **Phenotypes** |  |  |  |  |
| Heart failure n (%) | 8357 (7.2%) | 5336 (8.7%) | 377 (3.4%) | 183<br>(0.4%) |
| Aortic valve disease n (%) | 5526 (4.8%) | 3108 (5.0%) | 899 (8.2%) | 69 (0.2%) |
| Mitral valve disease n (%) | 9137 (7.9%) | 6564 (10.6%) | 2798<br>(25.5%) | 212<br>(0.5%) |
| Left ventricular<br>hypertrophy n (%) | 9909 (8.6%) | 3310 (5.4%) | 734 (6.7%) | 153<br>(0.4%) |
| **New-onset phenotypes** |  |  |  |  |
| N at risk | 97659 | 53046 | 8953 | 42014 |
| Median FU (yrs (iqr)) | 4.3 (2.5;5.0) | 3.7 (2.2;5.0) | 4.1 (3.0;5.0) | 5.0 (4.7;5.0) |
| Heart failure n (%) | 1980 (2.0%) | 1817 (3.4%) | 113 (1.3%) | 181 (0.4%) |
| Aortic valve disease n (%) | 1309 (1.3%) | 916 (1.7%) | 349 (3.9%) | 65 (0.2%) |
| Mitral valve disease n (%) | 711 (0.7%) | 687 (1.3%) | 292 (3.3%) | 65 (0.2%) |
| Left ventricular hypertrophy n (%) | 2374 (2.4%) | 1346 (2.5%) | 387 (4.3%) | 163 (0.4%) |
| Composite n (%) | 5177 (5.3%) | 4052 (7.6%) | 968 (10.8%) | 395 (0.9%) |

The cohort drawn from the community hospitals included 63,790 individuals who had a mean age of 61.1 ± 19.5 years, 36,088 (56.6%) were female, 41,401 (64.9%) self-reported as White, 7,760 (12.2%) as Black and 11,687 (18.3%) as Hispanic. Of these individuals, 32,021 (50.2%) had hypertension, 12,324 (19.3%) had diabetes, and 5,506 (8.6%) had heart failure. In the 11,005 individuals that were included from the outpatient clinic cohort, the mean age was 61.4 ± 15.0 years, and 6,241 (56.7%) were women. A total of 8,696 (79.0%) were White, 890 (8.1%) Black, and 928 (8.4%) Hispanic. Hypertension was present in 6,800 (61.8%), diabetes in 1,793 (16.3%), and heart failure in 408 (3.7%) individuals.

From the UK Biobank, the cohort consisted of 42,354 participants with a mean age of 64.1 ± 7.8 years, including 21,835 (51.6%) women, and were predominantly White (n=40,893, 96.6%). Hypertension was present in 6,269 (14.8%) of the individuals, diabetes in 1,340 (3.2%) and 225 (0.5%) had heart failure **(Table 1)**.

### Cross-sectional association of AI-ECG models with cardiovascular phenotypes

Across the 989 clinical phenotypes in the YNHH cohort, CVD-specific AI-ECG models were significantly more likely to be associated with phenotypes categorized as cardiovascular, compared with non-cardiovascular phenotypes, with ORs ranging from 4.41 (p = 2.26 × 10⁻¹^7^) for the AS model to 2.16 (p = 1.39 × 10⁻6) for the composite SHD model **(Figure 1, Supplementary Data 1)**. In contrast, the AI model trained to predict biological sex was not more likely to be associated with cardiovascular phenotypes but showed an increase in significant associations with genitourinary phenotypes (OR = 2.31; p = 1.34 × 10^−7^).

**Figure 1:**
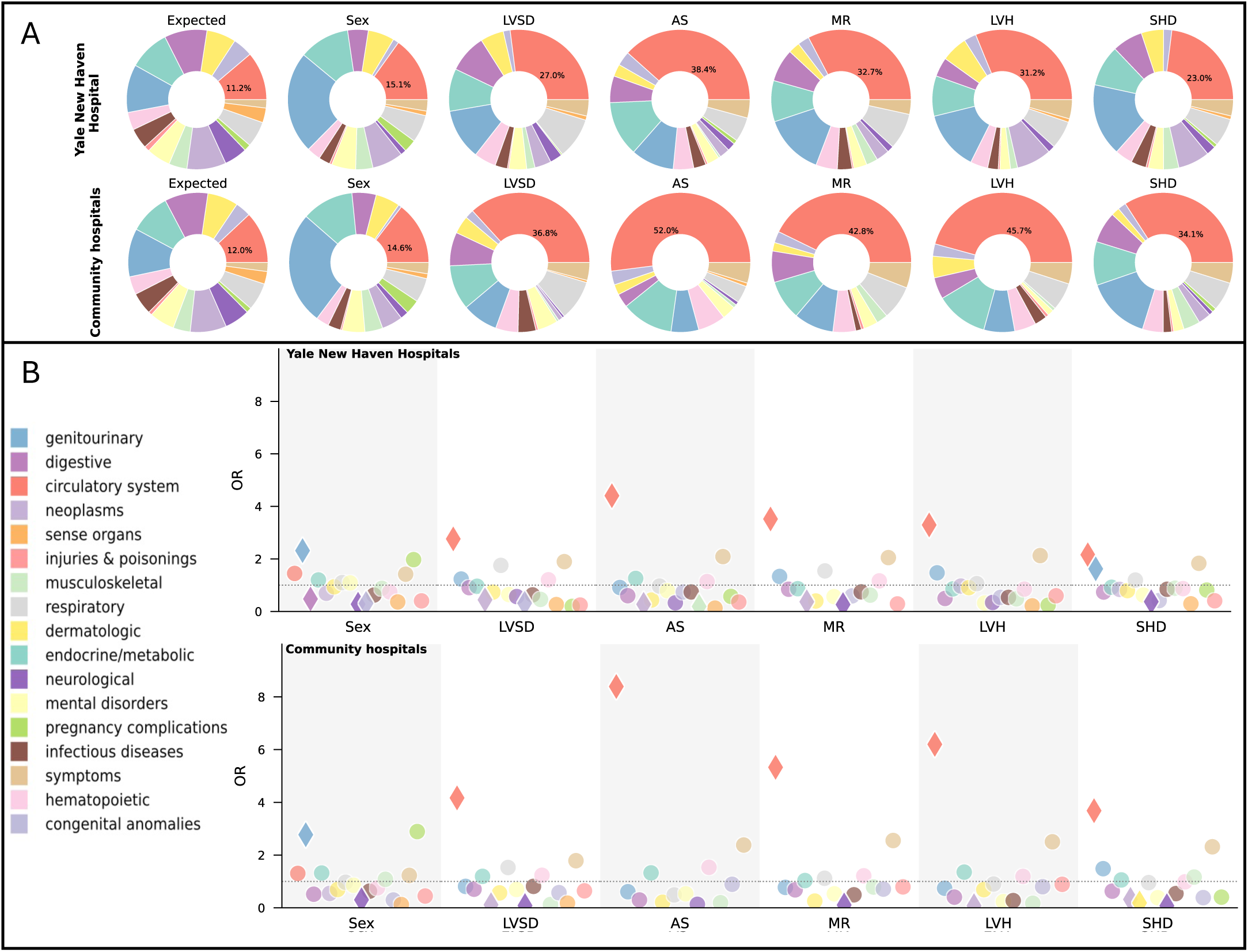
Overrepresentation of Associations with Cardiovascular Phenotypes by AI-ECG. **Panel A:** Distribution of significant associations per phenotype groups from the age sex adjusted serial logistic regression for each AI-ECG model in the Yale New Haven Health System and the four community hospitals. Each pie chart represents the proportion of significant phenotype associations across different organ systems or disease categories, as indicated by the color legend. The “Expected” distribution reflects the proportion of all tested phenotypes before applying significance thresholds. Across models, most associations relate to the circulatory system, except for the sex model. **Panel B:** The odds ratios (OR) for each phenotype category being significantly associated with the AI-ECG models compared with the expected distribution in the Yale New Haven Health System and the four community hospitals. Each marker represents the OR for a significant phenotype within a disease category. The y-axis represents the OR, while the x-axis corresponds to different AI-ECG models. Colors correspond to disease categories, diamonds indicate significant enrichment, and circles represent non-significant enrichment as shown as displayed in the legend. Abbreviations: AS, aortic stenosis; LVH, left ventricular hypertrophy; LVSD, left ventricular systolic dysfunction; MR, mitral regurgitation; SHD, structural heart disease; OR, odds ratio.

Similarly, among the available phenotypes from community hospitals (n=868), outpatient clinics (n=528), and the UK Biobank (n=687) model predictions for CVD specific AI-ECG models were consistently more likely to be associated with a broad range of cardiovascular phenotypes. In contrast, the AI-ECG model for biological sex classification did not show a cardiovascular specific phenotype association pattern **(Supplementary Data 1-4, Supplementary Figures 4-5)**. Additionally, while some associations between the cardiovascular disease models and non-cardiovascular phenotype groups were observed, they lacked consistency across models and cohorts **(Supplementary Data 1-4, Supplementary Figures 4-5)**.

### Off-target Phenotypic Associations of AI-ECG

We evaluated whether the AI-ECG models for detecting specific cardiovascular diseases were most strongly associated with their respective training labels. In the YNHH cohort, the LVSD model was strongly associated with its target phenotype, heart failure (OR per SD 2.91; 95% CI: 2.84-2.98). However, the model also showed strong associations with several off-target phenotypes, including primary/intrinsic cardiomyopathies (OR 3.58; 95% CI: 3.47-3.70), left bundle branch block (OR 3.11; 95% CI 2.96-3.26), and chronic ischemic heart disease (OR 2.93; 95% CI: 2.82-3.05) **(Figure 2B, Supplementary Data 5)**. This pattern was consistent across other AI-ECG models, each showing strong associations with a range of cardiovascular phenotypes including but not limited to its target phenotype **(Supplementary Figure 6)**.

**Figure 2:**
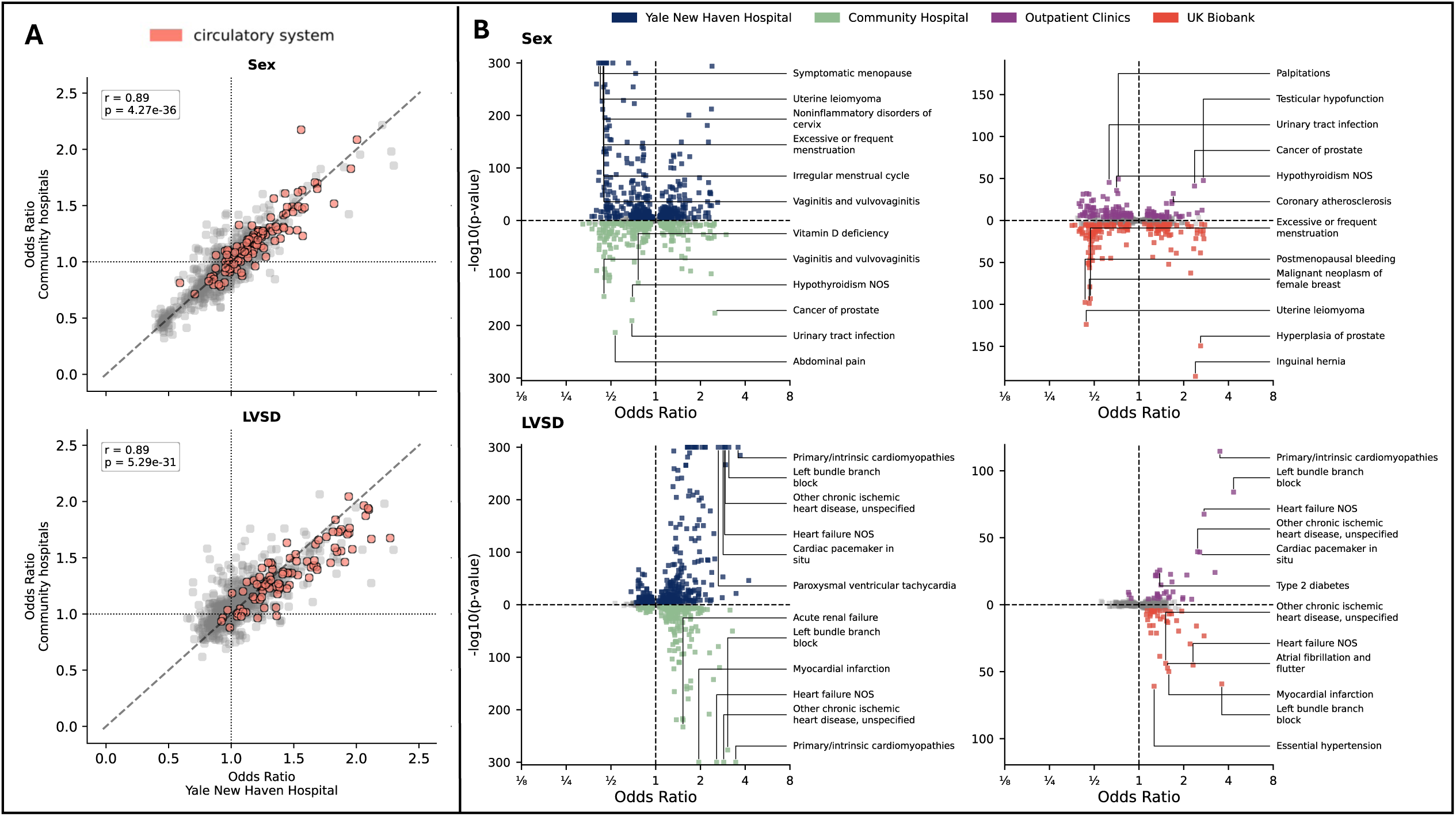
Associations between phenotypes and AI-ECG. **Panel A:** Scatter plots comparing odds ratios (ORs) between Yale New Haven Hospital (YNHH) and Community Hospitals for Sex and Mitral Regurgitation (MR). Data points corresponding to the circulatory system are highlighted in red. OR values are clipped at the 99th percentile to reduce the influence of outliers. The diagonal dashed line represents perfect agreement between the two cohorts. **Panel B:** Volcano plots displaying the associations between various phenotypes and AI-ECG model predictions across four cohorts: YNHH (blue), Community Hospitals (green), Outpatient Clinics (red), and UK Biobank (purple). The x-axis represents the odds ratios and are presented on the logarithmic scale, reflecting the effectsize and directionality of the association. The y-axis shows the –log₁₀(p-value) representing the significance of the association. Labeled points indicate the top 6 statistically significant associations. Due to maximal computational precision limits, p values are capped at 1×10^−300^. Abbreviations: AS, aortic stenosis; LVH, left ventricular hypertrophy; LVSD, left ventricular systolic dysfunction; MR, mitral regurgitation; SHD, structural heart disease; OR, odds ratio.

The sensitivity analysis restricting this cross-sectional analysis to individuals with progressively fewer concomitant diseases showed an attenuation of the effect sizes, but patterns similar to the primary analysis across all models **(Supplementary Figure 7)**. The specific examination of the AS model, LVH model and the HCM model showed that all 3 models have a high degree of overlap in the identification of each other’s target phenotypes, including the subphenotype of hypertrophic obstructive cardiomyopathy **(Supplementary Figure 8)**.

Associations between model outputs and phenotypes were consistent across the external validation cohorts **(Figure 2A; Supplementary Figures 9–11),** and off-target cardiovascular disease associations frequently exceeded those with the target phenotype. In the community hospitals, the LVSD model showed the strongest associations with primary/intrinsic cardiomyopathies (OR 3.45; 95% CI: 3.31-3.61), chronic ischemic heart disease (OR 2.87, 95% CI: 2.72-3.03) followed by its target heart failure (OR 2.57; 2.49-2.65) **(Supplementary Figure 6; Supplementary Data 6)**. Similar patterns were observed in the outpatient clinics and the UK Biobank.

For the AI-ECG model classifying biological sex, the most significant associations across cohorts were with sex-specific conditions **(Figure 2B; Supplementary Figure 6; Supplementary Data 5–8)**.

### Phenotypic Association Patterns of Disease-specific AI-ECG Models

In the YNHH cohort, the phenotype association patterns of the LVSD, MR, LVH, and SHD models were highly correlated, with Pearson correlation coefficients ranging from 0.88 to 0.99. In contrast, the AS model showed only moderate correlations with the other cardiovascular disease models (r = 0.61–0.86). **(Figure 3, Supplementary Figure 12)**. These patterns persisted when extending the analysis to include all cardiovascular phenotypes **(Supplementary Figure 13).**

**Figure 3:**
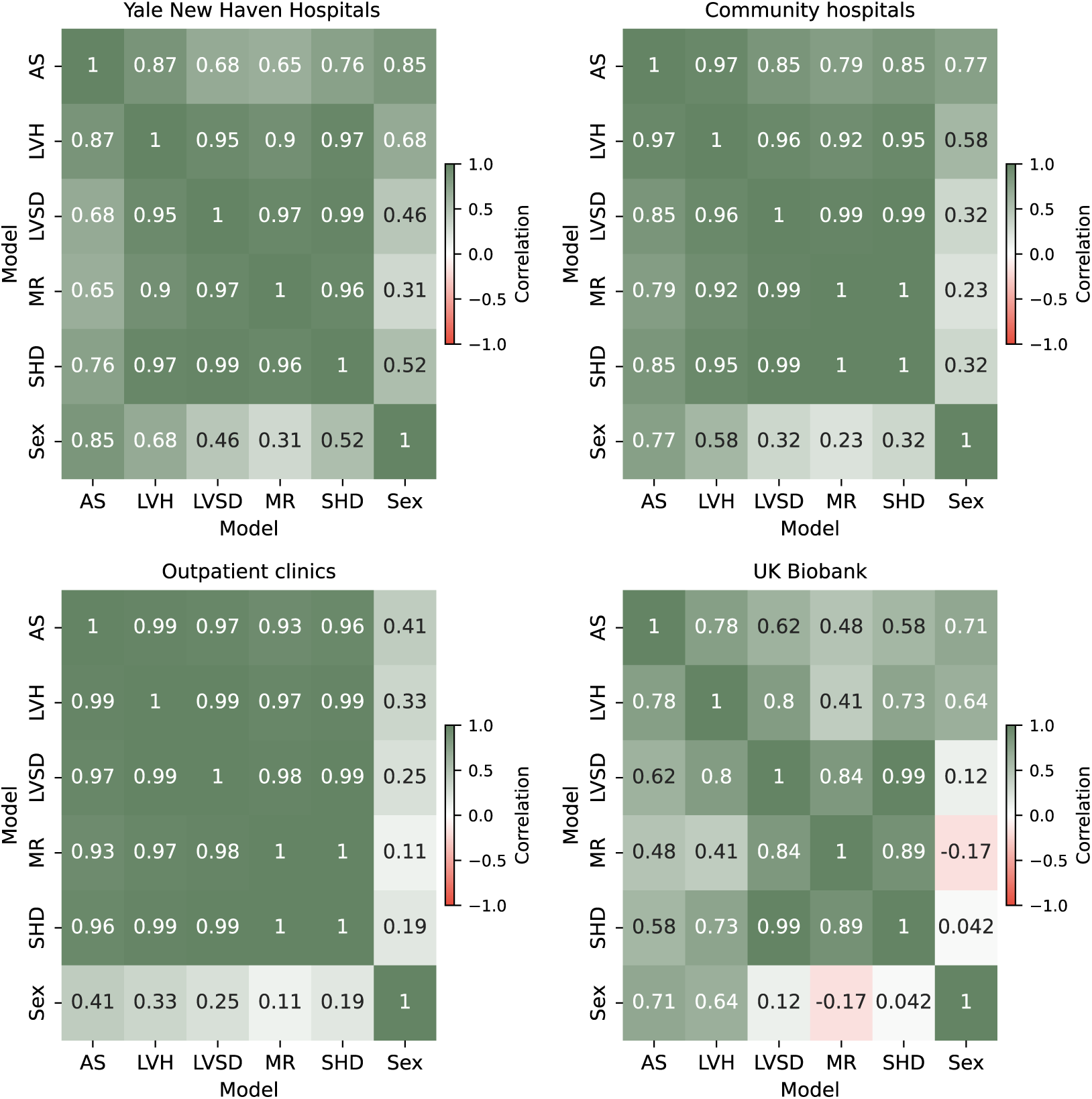
Correlation of on-target phenotype association profiles across AI-ECG models in four independent cohorts. These heatmaps show Pearson correlation coefficients between odds ratio (OR) profiles and AI-ECG models across all phenotypes that we defined as being the target for any of the models for each pair of AI-ECG models. Higher correlation coefficients indicate that models exhibit similar phenotypic association patterns across target cardiovascular conditions. Correlations are shown for four cohorts: Yale New Haven Hospitals, community hospitals, outpatient clinics, and the UK Biobank. Strong correlations were observed among cardiovascular disease (CVD) models, with the SHD model consistently showing high similarity with other disease-specific models. The model trained to predict biological sex showed consistently weaker correlation with cardiovascular models, particularly in the UK Biobank where inverse correlations were observed. AS: Aortic stenosis, LVH: Left ventricular hypertrophy, LVSD: Left ventricular systolic dysfunction, MR: Mitral regurgitation, SHD: Structural heart disease composite model, Sex: Biological sex prediction model.

### On– and Off-Target Cardiovascular Predictions with AI-ECG Models

After the exclusion of individuals with any of the target phenotypes before the ECG, 97659 individuals at risk had a median follow-up of 4.3 years (interquartile range (IQR), 2.5–5.0) in the YNHH cohort. During this period, 1,9580 individuals (2.0%) developed heart failure, 1,309 (1.3%) aortic valve disease, 711 (0.7%) mitral valve disease, and 2,374 (2.4%) LVH, collectively representing 5,177 individuals (5.3%) with an adverse cardiovascular outcome. The population characteristics, follow-up times, and incidence of key cardiovascular phenotypes for all the cohorts are detailed in **Table 1**.

Across models, heart failure consistently emerged as the top-ranked phenotype in the YNHH **(Figure 4; Supplementary Figure 14–16; Supplementary Data 11–13)**. The LVSD model was associated 2-fold elevated hazard for new onset HF (HR 2.09; 95% CI: 2.01–2.17), along with an elevated risk for a range of incident cardiovascular diseases, including aortic valve disease (HR 1.35; 95% CI: 1.28– 1.42), mitral valve disease (HR 1.67; 95% CI: 1.56–1.79), LVH (HR 1.69; 95% CI: 1.63–1.75) and composite SHD (HR 1.67; 95% CI: 1.63–1.72) for the composite of all these individual cardiovascular phenotypes.

**Figure 4:**
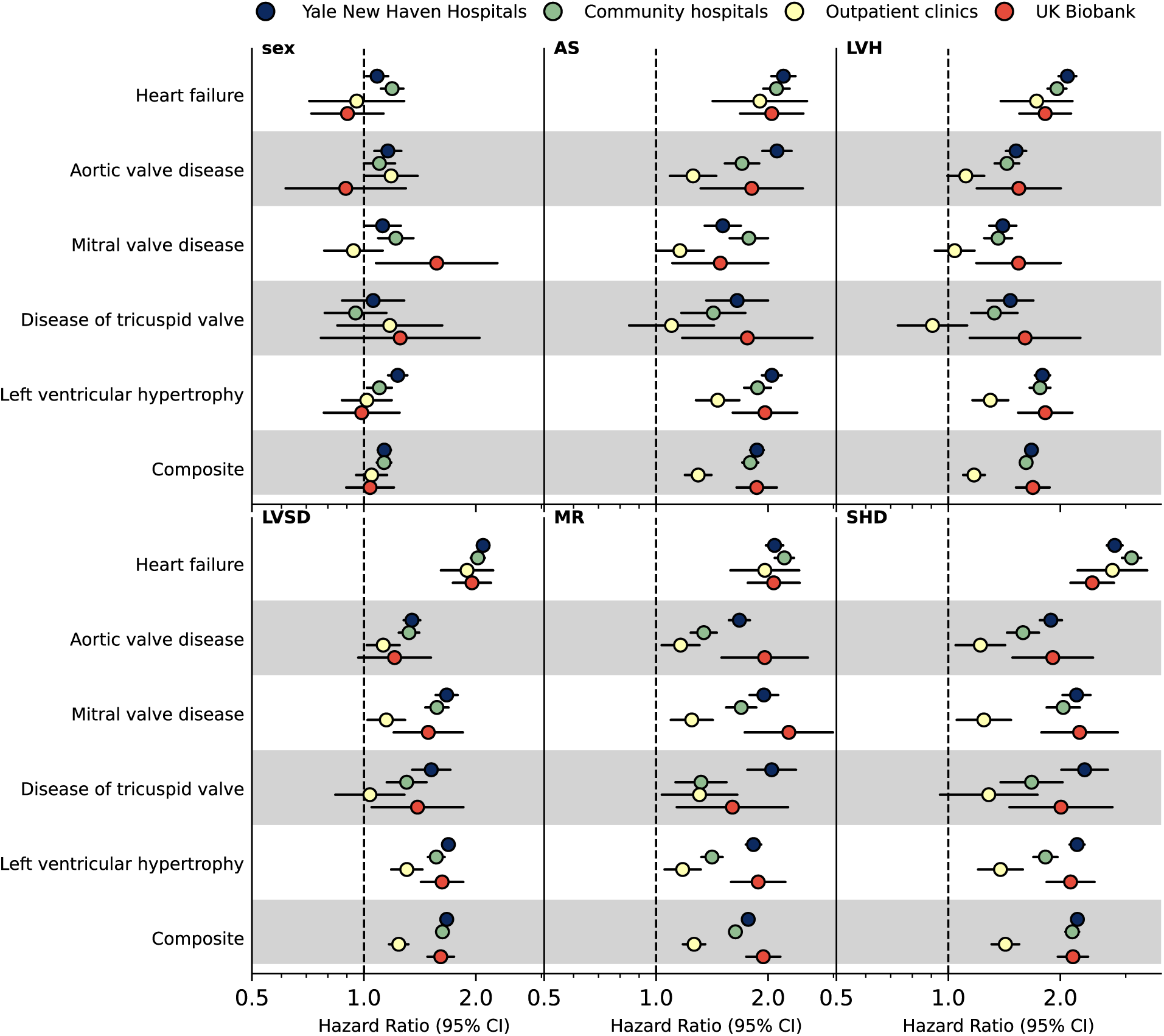
Forest plot displaying the results of the Cox Proportional Hazards models. Forest plot displaying the hazard ratios (HR) with 95% confidence intervals (CI) for various cardiovascular conditions across four cohorts: UK Biobank (UKB, red), Yale New Haven Hospital (YNHH, blue), Community Hospitals (green), and NEMG outpatient clinics (yellow). Each panel represents a different AI-ECG model prediction, including Sex, Mitral Regurgitation (MR), Left Ventricular Systolic Dysfunction (LVSD), Left Ventricular Hypertrophy (LVH), Aortic Stenosis (AS), and Structural Heart Disease (SHD). The x-axis represents the hazard ratio, with the vertical dashed line at HR = 1.0 indicating no association. The analysis was conducted using age– and sex-adjusted Cox regression models. Abbreviations: LVSD, Left Ventricular Systolic Dysfunction; LVH, Left Ventricular Hypertrophy; MR, Mitral Regurgitation; AS, Aortic Stenosis; SHD, Structural Heart Disease.

These findings were consistent across cohorts **(Figure 4; Supplementary Figures 14–16; Supplementary Data 11–13)**. In the community hospitals, all AI-ECG models were also significantly associated with the target phenotypes. In the outpatient clinics and the UK Biobank, associations were generally consistent but less uniformly significant **(Figure 4; Supplementary Figures 14–16)**. The AI-ECG model trained to predict biological sex was not significantly associated with any new-onset cardiovascular phenotype in YNHH, a finding that was consistent across all cohorts.

### Experimental Models for Non-Cardiovascular Disease Outcomes

Experimental Models for Non-Cardiovascular Disease Outcomes Across all cohorts, experimental AI-ECG models trained on non-cardiovascular outcomes were not significantly associated with clinical phenotypes, consistent with their role as negative controls. A limited number of associations were observed in the UK Biobank cohort, predominantly with cardiovascular conditions such as essential hypertension and coronary atherosclerosis. These findings were not replicated in other datasets **(Supplementary Figures 17–18)**.

## DISCUSSION

In this study of over 230,000 individuals from four distinct clinical and community-based cohort studies across the US and UK, we demonstrate that a range of AI-ECG models developed to identify specific structural cardiac disorders have non-specific associations with a range of cardiovascular conditions. These models used standard 12-lead ECG images as input, with prior validation in multinational cohorts, and assessments confirming consistency with signal-based models in prior reports.^3,19^ While these models consistently showed stronger associations with cardiovascular phenotypes, these associations were notable across both the target and non-target conditions, suggesting that they function better as general cardiovascular screening tools than as condition-specific diagnostic modalities. This pattern was consistently observed across cohorts with prevalent and frequently coexisting cardiovascular conditions, in sensitivity analyses with limited multimorbidity, and in a healthier population initially free of structural heart disease. Moreover, these tools represent digital biomarkers of future cardiovascular disease in individuals without overt cardiac dysfunction. The strongest association was found between these AI-ECG tools and the risk of new-onset HF, rather than the risk of future development of the target conditions. Notably, the ensemble model trained to identify a composite of structural heart diseases exhibited broader and stronger associations than models trained on individual conditions.

Our study presents essential insights to interpret the evolving field of AI-ECG-based cardiovascular screening. Currently, there has been a preponderance of AI-ECG models developed for label specific classification tasks for a variety of cardiovascular diseases.^1,3,11,33,36–38^ This approach has led to numerous AI-ECG models demonstrating the strong discriminatory performance for the detection of these specific conditions.^17,39,40^ However, our head-to-head evaluation of AI-ECG models revealed that the limited selectivity of these models hinders their utility for disease-specific screening.^41^ This may be a result of limited understanding of what an abnormal prediction on AI-ECG models may represent.^8,33^ This is particularly relevant as the exclusion of the target condition in those with abnormal AI-ECG predictions may still represent predisposition for a range of other cardiovascular conditions.^42^ Before the widespread clinical implementation of AI-ECG, it is essential to inform clinicians what these positive screens across different AI-ECG models may mean for patients and to understand whether an algorithm’s output reflects a precise signal linked to its intended target condition or whether it captures broader, non-specific disease signatures.

In contrast to the prevailing disease-specific paradigm, our findings suggest that AI-ECG models, despite being trained to detect specific cardiovascular disease labels, capture a highly correlated, broader physiological signature rather than distinct disease-specific features, thereby reframing the role of AI-ECG from that of binary diagnostic classifiers to biomarkers of cardiovascular vulnerability. This fits in the growing recognition that ECGs encode rich, latent information spanning a wide range of both cardiovascular and non-cardiovascular conditions.^43,44^ While current approaches fine-tune AI-ECG models to use this information for the abstraction of disease-specific ECG signatures, our data revealed that this narrow strategy offers limited clinical utility.^1,45^

To examine these shared signatures, we evaluated a broad number of phenotypic associations across models. For example, left bundle branch block, a recognizable conduction abnormality, emerged consistently as a major association across multiple model outputs and is clinically associated with a range of cardiovascular diseases.^46,47^ Other frequently associated traits included hypertension, diabetes, and renal dysfunction, conditions known to drive systemic and cardiac remodeling not limited to a specific disease, further substantiating that AI-ECG models may be detecting subtle rhythm and conduction disturbances linked to generalized cardiovascular risk, or confounders of the target disease label used in model development.^46,47^

Despite the limited utility of disease-specific models, the choice of training label substantially shaped performance and clinical applicability. In our experiments, a model trained on a well-defined composite endpoint of structural heart diseases produced broader, more robust associations than models trained on single disease labels. Conversely, a biological sex classification model revealed distinct non-cardiovascular signatures, and models trained on random or ECG-unrelated labels yielded no meaningful patterns. These results suggest that the future development of AI-ECG algorithms would be most optimized when focused on detecting well-defined composite cardiovascular outcomes.

These findings carry important implications for the future development, regulatory approval, and subsequent implementation and reimbursement of AI-ECG models. An abnormal AI-ECG-based risk profile should prompt a comprehensive cardiovascular evaluation, including echocardiography, to uncover structural pathology beyond the model’s target. Moreover, high scores independently predict future cardiovascular events even in the absence of overt disease, underscoring AI-ECG’s role as a prognostic biomarker. Incorporating these continuous risk outputs into clinical workflows can guide early preventive strategies, such as lipid-lowering therapies, intensified blood pressure control, and lifestyle modification.^48^ In future research, developers of AI-ECG tools could specifically target the development of models that differentiate between key subphenotypes, which may improve their role in scenarios where this would be clinically meaningful. This strategy has been effective in other AI domains, including AI-based echocardiography.^49,50^

These findings may also have important implications for the current regulatory and reimbursement frameworks for AI-ECG. Recently, the FDA has granted 510(k) clearance to algorithms for models detecting hypertrophic cardiomyopathy and LVSD, and CMS has introduced a reimbursable code for AI-ECG interpretation. ^51^ However, prevailing evaluation frameworks remain anchored in disease-specific, binary classification, which can lead to separate approval and coverage pathways for multiple algorithms.^52^ Given our observation that models trained on distinct conditions have a highly correlated phenotypic pattern, it may be important to evaluate whether such models truly function as disease-specific classifiers, and shift the evaluation of AI-ECG to using a single AI-ECG model as a broad cardiovascular risk classifier, rather than relying on multiple disease-specific tools.

Our study should be interpreted considering the following potential limitations. Phenotype classification relied on ICD diagnostic codes, which are subject to misclassification and variability across clinical settings, potentially affecting the accuracy of our outcome definitions. Moreover, the use of diagnostic codes may result in the potential underrepresentation of certain phenotypes. This is particularly important in the UK Biobank, where only inpatient codes are available, resulting in the low phenotype counts for certain phenotypes, such as LVH. However, the consistent results across distinct external validation populations suggest the generalizability of the findings despite differences in coding practices. Furthermore, while there may be indication bias in ECG acquisition within the clinical cohorts where ECGs are typically performed for specific clinical reasons, potentially leading to a higher prevalence of CVD with the general population, the UK Biobank cohort consists of research participants who underwent ECGs as part of a structured protocol, reducing the likelihood of such bias. Additionally, PheWAS analyses involve testing a large number of phenotypes, which increases the risk of false positives. However, we applied strict multiple testing correction and replication in independent datasets.^3,19,53^ Moreover, AI-ECG models were trained on historical datasets with structured outcome labels, which may not capture the full nuance of real-world clinical practice or longitudinal disease progression. Since all models utilized in this study are based on a single architecture and trained in the same population, caution is warranted when extrapolating our findings to AI-ECG models developed using different architectures or trained in distinct populations. However, image-based models, including those used in this study, have been extensively validated and have demonstrated similar performance to signal-based models in head-to-head comparison, with high correlation and comparable accuracy across multiple studies and independent datasets.^3,19,33,54,55^ The study, however, suggests the role of such selectivity assessments across different models before recommending their use as single disease diagnostic agents. Finally, while our findings suggest predictive utility, they remain observational. Prospective validation of a composite AI-ECG model in pragmatic trials will be critical to translate these findings into clinical practice.

## Conclusion

Despite being developed for the detection of specific cardiovascular conditions, AI-ECG models both detect the presence as well as predict the future development of a broad range of cardiovascular diseases with similar propensity. This challenges their role as distinct binary diagnostic tools and instead suggests the clinical utility of AI-ECG as a broader cardiovascular biomarker.

## Data Availability

Due to the inclusion of patient-level data, the dataset cannot be publicly shared; however, researchers may contact the corresponding author to request access for academic purposes.

## Non-standard Abbreviations and Acronyms

AI: Artificial Intelligence
AI-ECG: Artificial Intelligence-enhanced Electrocardiogram
AS: Aortic Stenosis
AUROC: Area Under the Receiver Operating Characteristic Curve
CNN: Convolutional Neural Network
CVD: Cardiovascular Disease
EHR: Electronic Health Record
ECG: Electrocardiogram
FDA: Food and Drug Administration
ICD: International Classification of Diseases
LVEF: Left Ventricular Ejection Fraction
LVH: Left Ventricular Hypertrophy
LVSD: Left Ventricular Systolic Dysfunction
MR: Mitral Regurgitation
HCM: Hypertrophic cardiomyopathy
HOCM: Hypertrophic obstructive cardiomyopathy
NHS: National Health Service
PheWAS: Phenome-Wide Association Study
SHD: Structural Heart Disease
UKB: UK Biobank
YNHH: Yale New Haven Hospital
YNHHS: Yale New Haven Health System

## Supplemental Material

Supplemental Materials

Supplementary Tables 1–5

Supplementary Figures 1-18

Supplementary Data 1-12

## References

1. Siontis KC, Noseworthy PA, Attia ZI, Friedman PA. Artificial intelligence-enhanced electrocardiography in cardiovascular disease management. Nat Rev Cardiol. 2021;18:465–478.

2. Somani S, Russak AJ, Richter F, Zhao S, Vaid A, Chaudhry F, De Freitas JK, Naik N, Miotto R, Nadkarni GN, Narula J, Argulian E, Glicksberg BS. Deep learning and the electrocardiogram: review of the current state-of-the-art. Europace. 2021;23:1179–1191.

3. Sangha V, Nargesi AA, Dhingra LS, Khunte A, Mortazavi BJ, Ribeiro AH, Banina E, Adeola O, Garg N, Brandt CA, Miller EJ, Ribeiro ALP, Velazquez EJ, Giatti L, Barreto SM, Foppa M, Yuan N, Ouyang D, Krumholz HM, Khera R. Detection of left ventricular systolic dysfunction from electrocardiographic images. Circulation. 2023;148:765–777.

4. Tsai D-J, Lou Y-S, Lin C-S, Fang W-H, Lee C-C, Ho C-L, Wang C-H, Lin C. Mortality risk prediction of the electrocardiogram as an informative indicator of cardiovascular diseases. Digit Health. 2023;9:20552076231187250.

5. Attia ZI, Kapa S, Lopez-Jimenez F, McKie PM, Ladewig DJ, Satam G, Pellikka PA, Enriquez-Sarano M, Noseworthy PA, Munger TM, Asirvatham SJ, Scott CG, Carter RE, Friedman PA. Screening for cardiac contractile dysfunction using an artificial intelligence-enabled electrocardiogram. Nat Med. 2019;25:70–74.

6. Kwon J-M, Kim K-H, Akkus Z, Jeon K-H, Park J, Oh B-H. Artificial intelligence for detecting mitral regurgitation using electrocardiography. J Electrocardiol. 2020;59:151–157.

7. Kwon J-M, Jeon K-H, Kim HM, Kim MJ, Lim SM, Kim K-H, Song PS, Park J, Choi RK, Oh B-H. Comparing the performance of artificial intelligence and conventional diagnosis criteria for detecting left ventricular hypertrophy using electrocardiography. Europace. 2020;22:412–419.

8. Dhingra LS, Aminorroaya A, Sangha V, Pedroso AF, Asselbergs FW, Brant LCC, Barreto SM, Ribeiro ALP, Krumholz HM, Oikonomou EK, Khera R. Heart failure risk stratification using artificial intelligence applied to electrocardiogram images: a multinational study. Eur Heart J. 2025;46:1044–1053.

9. Noseworthy PA, Attia ZI, Behnken EM, Giblon RE, Bews KA, Liu S, Gosse TA, Linn ZD, Deng Y, Yin J, Gersh BJ, Graff-Radford J, Rabinstein AA, Siontis KC, Friedman PA, Yao X. Artificial intelligence-guided screening for atrial fibrillation using electrocardiogram during sinus rhythm: a prospective non-randomised interventional trial. Lancet. 2022;400:1206–1212.

10. Sau A, Pastika L, Sieliwonczyk E, Patlatzoglou K, Ribeiro AH, McGurk KA, Zeidaabadi B, Zhang H, Macierzanka K, Mandic D, Sabino E, Giatti L, Barreto SM, Camelo L do V, Tzoulaki I, O’Regan DP, Peters NS, Ware JS, Ribeiro ALP, Kramer DB, Waks JW, Ng FS. Artificial intelligence-enabled electrocardiogram for mortality and cardiovascular risk estimation: a model development and validation study. Lancet Digit Health. 2024;6:e791–e802.

11. Sau A, Barker J, Pastika L, Sieliwonczyk E, Patlatzoglou K, McGurk KA, Peters NS, O’Regan DP, Ware JS, Kramer DB, Waks JW, Ng FS. Artificial intelligence-enhanced electrocardiography for prediction of incident hypertension. JAMA Cardiol. 2025;10:214–223.

12. Anumana ECG-AI LEF [Internet]. [cited 2025 Feb 11];Available from: https://anumana.ai/ecg-ai-lef

13. Kumar V, Venkataraman R, Aljaroudi W, Osorio J, Heo J, Iskandrian AE, Hage FG. Implications of left bundle branch block in patient treatment. Am J Cardiol. 2013;111:291–300.

14. Sahranavard T, Alimi R, Arabkhazaei J, Nasrabadi M, Alavi Dana SMM, Gholami Y, Izadi-Moud A, Esmaily H, Ebrahimi M, Ferns GA, Moohebati M, Saffar Soflaei S, Ghayour Mobarhan M. Association of major and minor ECG abnormalities with traditional cardiovascular risk factors in the general population: a large scale study. Sci Rep. 2024;14:11289.

15. Jørgensen PG, Jensen JS, Marott JL, Jensen GB, Appleyard M, Mogelvang R. Electrocardiographic changes improve risk prediction in asymptomatic persons age 65 years or above without cardiovascular disease. J Am Coll Cardiol. 2014;64:898–906.

16. Hari KJ, Singleton MJ, Ahmad MI, Soliman EZ. Relation of minor electrocardiographic abnormalities to cardiovascular mortality. Am J Cardiol. 2019;123:1443–1447.

17. Croon PM, Pedroso AF, Khera R. The emerging role of AI in transforming cardiovascular care. Future Cardiol. 2025;1–4.

18. Zoghbi WA, Adams D, Bonow RO, Enriquez-Sarano M, Foster E, Grayburn PA, Hahn RT, Han Y, Hung J, Lang RM, Little SH, Shah DJ, Shernan S, Thavendiranathan P, Thomas JD, Weissman NJ. Recommendations for noninvasive evaluation of native valvular regurgitation: A report from the American society of echocardiography developed in collaboration with the society for cardiovascular magnetic resonance. J Am Soc Echocardiogr. 2017;30:303–371.

19. Dhingra LS, Aminorroaya A, Sangha V, Pedroso AF, Shankar SV, Coppi A, Foppa M, Brant LCC, Barreto SM, Ribeiro ALP, Krumholz HM, Oikonomou EK, Khera R. Ensemble deep learning algorithm for structural heart disease screening using electrocardiographic images. J Am Coll Cardiol. 2025;85:1302– 1313.

20. Sangha V, Khunte A, Holste G, Mortazavi BJ, Wang Z, Oikonomou EK, Khera R. Biometric contrastive learning for data-efficient deep learning from electrocardiographic images. J Am Med Inform Assoc. 2024;31:855–865.

21. Kossaify A, Nasr M. Diastolic dysfunction and the new recommendations for echocardiographic assessment of left ventricular diastolic function: Summary of guidelines and novelties in diagnosis and grading. J Diagn Med Sonogr. 2019;35:317–325.

22. Attia ZI, Friedman PA, Noseworthy PA, Lopez-Jimenez F, Ladewig DJ, Satam G, Pellikka PA, Munger TM, Asirvatham SJ, Scott CG, Carter RE, Kapa S. Age and sex estimation using artificial intelligence from standard 12-lead ECGs. Circ Arrhythm Electrophysiol. 2019;12:e007284.

23. Denny JC, Ritchie MD, Basford MA, Pulley JM, Bastarache L, Brown-Gentry K, Wang D, Masys DR, Roden DM, Crawford DC. PheWAS: demonstrating the feasibility of a phenome-wide scan to discover gene-disease associations. Bioinformatics. 2010;26:1205–1210.

24. Boland MR, Alur-Gupta S, Levine L, Gabriel P, Gonzalez-Hernandez G. Disease associations depend on visit type: results from a visit-wide association study. BioData Min. 2019;12:15.

25. Fan M, Li N, Huang L, Chen C, Dong X, Gao W. Exploring potential drug targets in multiple cardiovascular diseases: A study based on proteome-wide Mendelian randomization and colocalization analysis. Cardiovasc Ther. 2025;2025:5711316.

26. Wan NC, Grabowska ME, Kerchberger VE, Wei W-Q. Exploring beyond diagnoses in electronic health records to improve discovery: a review of the phenome-wide association study. JAMIA Open. 2025;8:ooaf006.

27. Denny JC, Bastarache L, Ritchie MD, Carroll RJ, Zink R, Mosley JD, Field JR, Pulley JM, Ramirez AH, Bowton E, Basford MA, Carrell DS, Peissig PL, Kho AN, Pacheco JA, Rasmussen LV, Crosslin DR, Crane PK, Pathak J, Bielinski SJ, Pendergrass SA, Xu H, Hindorff LA, Li R, Manolio TA, Chute CG, Chisholm RL, Larson EB, Jarvik GP, Brilliant MH, McCarty CA, Kullo IJ, Haines JL, Crawford DC, Masys DR, Roden DM. Systematic comparison of phenome-wide association study of electronic medical record data and genome-wide association study data. Nat Biotechnol. 2013;31:1102–1110.

28. PheWAS Resources [Internet]. [cited 2025 Mar 21];Available from: https://phewascatalog.org/phewas/#phe12

29. Ritchie MD, Denny JC, Crawford DC, Ramirez AH, Weiner JB, Pulley JM, Basford MA, Brown-Gentry K, Balser JR, Masys DR, Haines JL, Roden DM. Robust replication of genotype-phenotype associations across multiple diseases in an electronic medical record. Am J Hum Genet. 2010;86:560–572.

30. Wu P, Gifford A, Meng X, Li X, Campbell H, Varley T, Zhao J, Carroll R, Bastarache L, Denny JC, Theodoratou E, Wei W-Q. Mapping ICD-10 and ICD-10-CM codes to phecodes: Workflow development and initial evaluation. JMIR Med Inform. 2019;7:e14325.

31. Wei W-Q, Bastarache LA, Carroll RJ, Marlo JE, Osterman TJ, Gamazon ER, Cox NJ, Roden DM, Denny JC. Evaluating phecodes, clinical classification software, and ICD-9-CM codes for phenome-wide association studies in the electronic health record. PLoS One. 2017;12:e0175508.

32. Akbilgic O, Butler L, Karabayir I, Chang PP, Kitzman DW, Alonso A, Chen LY, Soliman EZ. ECG-AI: electrocardiographic artificial intelligence model for prediction of heart failure. Eur Heart J Digit Health. 2021;2:626–634.

33. Sangha V, Dhingra LS, Aminorroaya A, Croon PM, Sikand NV, Sen S, Martinez MW, Maron MS, Krumholz HM, Asselbergs FW, Oikonomou EK, Khera R. Identification of hypertrophic cardiomyopathy on electrocardiographic images with deep learning. Nat Cardiovasc Res. 2025;1–10.

34. Carroll RJ, Bastarache L, Denny JC. R PheWAS: data analysis and plotting tools for phenome-wide association studies in the R environment. Bioinformatics. 2014;30:2375–2376.

35. PheWAS: The PheWAS R package [Internet]. Github; [cited 2025 Feb 12]. Available from: https://github.com/PheWAS/PheWAS

36. Yao X, Rushlow DR, Inselman JW, McCoy RG, Thacher TD, Behnken EM, Bernard ME, Rosas SL, Akfaly A, Misra A, Molling PE, Krien JS, Foss RM, Barry BA, Siontis KC, Kapa S, Pellikka PA, Lopez-Jimenez F, Attia ZI, Shah ND, Friedman PA, Noseworthy PA. Artificial intelligence-enabled electrocardiograms for identification of patients with low ejection fraction: a pragmatic, randomized clinical trial. Nat Med. 2021;27:815–819.

37. Oikonomou EK, Sangha V, Shankar SV, Coppi A, Krumholz HM, Nasir K, Miller EJ, Gallegos-Kattan C, Al-Kindi S, Khera R. Tracking the pre-clinical progression of transthyretin amyloid cardiomyopathy using artificial intelligence-enabled electrocardiography and echocardiography. medRxiv [Internet]. 2024 [cited 2024 Oct 9];Available from: https://pubmed.ncbi.nlm.nih.gov/39252891/

38. Kwon J-M, Kim K-H, Jo Y-Y, Jung M-S, Cho Y-H, Shin J-H, Lee Y-J, Ban J-H, Lee SY, Park J, Oh B-H. Artificial intelligence assessment for early detection and prediction of renal impairment using electrocardiography. Int Urol Nephrol. 2022;54:2733–2744.

39. Palermi S, Vecchiato M, Ng FS, Attia Z, Cho Y, Anselmino M, De Ferrari GM, Saglietto A, International AI-ECG Working Group. Artificial intelligence and the electrocardiogram: A modern renaissance. Eur J Intern Med [Internet]. 2025 [cited 2025 Jun 17];Available from: 10.1016/j.ejim.2025.04.036

40. Mossavarali S, Vaezi A, Gholami Z, Molaei A, Yekaninejad MS, Asselbergs FW, Shafiee A. Determinants of artificial intelligence electrocardiogram-derived age and its association with cardiovascular events and mortality: a systematic review and meta-analysis. NPJ Digit Med. 2025;8:322.

41. Lampert J, Bhatt DL, Vaid A, Kon K, Feinman J, Jou S, Kauffman J, Nadkarni G, Reddy VY. Calibration of ECG-based deep-learning algorithm scores for patients flagged as high risk for hypertrophic cardiomyopathy. NEJM AI [Internet]. 2025 [cited 2025 May 5];2. Available from: 10.1056/aioa2400421

42. Yoon M, You SC. Implementing ECG-AI in clinical care: From pixel to practice. J Am Coll Cardiol. 2025;85:1314–1316.

43. Friedman SF, Khurshid S, Venn RA, Wang X, Diamant N, Di Achille P, Weng L-C, Choi SH, Reeder C, Pirruccello JP, Singh P, Lau ES, Philippakis A, Anderson CD, Maddah M, Batra P, Ellinor PT, Ho JE, Lubitz SA. Unsupervised deep learning of electrocardiograms enables scalable human disease profiling. NPJ Digit Med. 2025;8:23.

44. Hughes JW, Theurer J, Vukadinovic M, Rogers AJ, Somani S, Kang G, Ghazizadeh Z, O’Sullivan JW, Jain SS, Gomes B, Salerno M, Ashley E, Zou JY, Perez MV, Ouyang D. A deep learning phenome wide association study of the electrocardiogram. Eur Heart J Digit Health. 2025;ztaf047.

45. Muzammil MA, Javid S, Afridi AK, Siddineni R, Shahabi M, Haseeb M, Fariha FNU, Kumar S, Zaveri S, Nashwan AJ. Artificial intelligence-enhanced electrocardiography for accurate diagnosis and management of cardiovascular diseases. J Electrocardiol. 2024;83:30–40.

46. Fuchs FD, Whelton PK. High blood pressure and cardiovascular disease. Hypertension. 2020;75:285–292.

47. Deferrari G, Cipriani A, La Porta E. Renal dysfunction in cardiovascular diseases and its consequences. J Nephrol. 2021;34:137–153.

48. Antoniades C, Chan K. Using artificial intelligence to spot heart failure from ECGs: is it prime time? Eur Heart J. 2025;46:1054–1056.

49. Slivnick JA, Hawkes W, Oliveira J, Woodward G, Akerman A, Gomez A, Hamza I, Desai VK, Barrett-O’Keefe Z, Grogan M, Dispenzieri A, Scott CG, Davison HN, Cotella J, Maurer M, Helmke S, Scherrer-Crosbie M, Soltani M, Goyal A, Zareba KM, Cheng RK, Kirkpatrick JN, Kitano T, Takeuchi M, Tiemi Hotta V, Campos Vieira ML, Elissamburu P, Ronderos RE, Prado A, Koutroumpakis E, Deswal A, Pursnani A, Sarswat N, Patel AR, Addetia K, Ruberg FL, Randazzo M, Asch FM, O’Driscoll J, Al-Roub N, Strom JB, Kidd L, Cuddy S, Upton R, Lang RM, Pellikka PA. Cardiac amyloidosis detection from a single echocardiographic video clip: a novel artificial intelligence-based screening tool. Eur Heart J [Internet]. 2025; Available from: 10.1093/eurheartj/ehaf387

50. Vrudhula A, Stern L, Cheng PC, Ricchiuto P, Daluwatte C, Witteles R, Patel J, Ouyang D. Impact of case and control selection on training artificial intelligence screening of cardiac amyloidosis. JACC Adv. 2024;3:100998.

51. Babic B, Glenn Cohen I, Stern AD, Li Y, Ouellet M. A general framework for governing marketed AI/ML medical devices. NPJ Digit Med. 2025;8:328.

52. Windecker D, Baj G, Shiri I, Kazaj PM, Kaesmacher J, Gräni C, Siontis GCM. Generalizability of FDA-approved AI-enabled medical devices for clinical use. JAMA Netw Open. 2025;8:e258052.

53. Bastarache L, Denny JC, Roden DM. Phenome-wide association studies. JAMA. 2022;327:75–76.

54. Croon PM, Boonstra MJ, Allaart CP, Arends BKO, Dhingra LS, Huang Y-C, Mast T, Khera R, Kuo C-F, Kwon J-M, Lee H-S, Lee MS, van de Leur RR, Liu Z-Y, Oikonomou EK, Selder JL, Winter MM, Asselbergs FW. AI-ECG for LVSD detection: a systematic review and first-in-kind multinational head-to-head comparison [Internet]. medRxiv. 2025; Available from: 10.1101/2025.07.08.25331129

55. Sau A, Zeidaabadi B, Patlatzoglou K, Pastika L, Ribeiro AH, Sabino E, Peters NS, Ribeiro ALP, Kramer DB, Waks JW, Ng FS. A comparison of artificial intelligence-enhanced electrocardiography approaches for prediction of time-to-mortality using electrocardiogram images. Eur Heart J Digit Health. 2024;ztae090.

