## Supplementary Materials for "Phenotypic Selectivity of Artificial Intelligence-enhanced Electrocardiography in Cardiovascular Diagnosis and Risk Prediction"

Philip M. Croon … Rohan Kera et al.

**Supplementary Materials**

### **Supplementary tables**

**Supplementary table 1:** Performance of AI-ECG for male sex

| **Cohort** | **Mean AUC** | **Lower CI** | **Upper CI** |
| --- | --- | --- | --- |
| **Yale New Haven Hospitals** | 0.917 | 0.916 | 0.919 |
| **Community Hospitals** | 0.902 | 0.899 | 0.904 |
| **Outpatient Clinics** | 0.945 | 0.943 | 0.948 |
| **UK Biobank** | 0.929 | 0.927 | 0.932 |

NB. Internal (Yale New Haven Health) and external (Community Hospitals, Outpatient Clinics, Uk Biobank) validation of the experimental male sex model. Abbreviations: AUC: Area under the Receiver Operating Curve, CI: Confidence Interval, UK: United Kingdom.

**Supplementary table 2:** ICD codes used for experimental non-cardiovascular disease phenotype models

| **Phenotype** | **ICD codes** |
| --- | --- |
| **Fracture of the lower leg** | ‘S82’ |
| **Dermatophytosis** | ‘B35’ |
| **Bitten by Dog** | ‘W54’ |
| **Viral Respiratory Infection** | ‘J09’, ’J10’, ‘J11’, ‘J12’, ‘J20’ |
| **Transport Accident** | ‘V00’ to ‘V79’ |

**Supplementary table 3:** Results of the experimental AI-ECG models.

| **Variable** | **n (%)** | **AUC** |
| --- | --- | --- |
| **Even month** | 132305 (49.6%) | 0.50 |
| **Fracture of the lower leg** | 3922 (1.5%) | 0.50 |
| **Dermatophytosis** | 14356 (5.4%) | 0.50 |
| **Bitten by dog** | 533 (0.2%) | 0.52 |
| **Viral respiratory infection** | 15251 (5.7%) | 0.50 |
| **Headache** | 30569 (11.5%) | 0.50 |
| **Transport accident** | 5434 (2.0%) | 0.51 |

**Supplementary table 4:** Diagnostic codes used for the definition of baseline characteristics

| **Condition** | **ICD-10 CM** |
| --- | --- |
| Heart failure | I110', 'I130', 'I132', 'I50', 'I500', 'I501', 'I509', 'Z9581', 'I0981' |
| Acute myocardial infarction | 'I21', 'I22', 'I23', 'I240', 'I248', 'I249' |
| Ischemic heart disease | I20', 'I200', 'I208', 'I209', 'I21', 'I210', 'I211', 'I212', 'I213',  'I214', 'I219', 'I21X', 'I22', 'I220', 'I221', 'I228', 'I229', 'I23',  'I230', 'I231', 'I232', 'I233', 'I234', 'I235', 'I236', 'I238', 'I24',  'I240', 'I241', 'I248', 'I249', 'I25', 'I250', 'I251', 'I252', 'I255',  'I256', 'I258', 'I259' |
| Stroke | G45', 'G450', 'G451', 'G452', 'G453', 'G454', 'G458', 'G459', 'I63',  'I630', 'I631', 'I632', 'I633', 'I634', 'I635', 'I638', 'I639', 'I64',  'I65', 'I650', 'I651', 'I652', 'I653', 'I658', 'I659', 'I66', 'I660',  'I661', 'I662', 'I663', 'I664', 'I668', 'I669', 'I672', 'I693', 'I694' |
| Hypertension | I10', 'I11', 'I110', 'I119', 'I12', 'I120', 'I129', 'I13', 'I130', 'I131',  'I132', 'I139', 'I674', 'O10', 'O100', 'O101', 'O102', 'O103', 'O109', 'O11' |
| Diabetes | E10', 'E100', 'E101', 'E102', 'E103', 'E104', 'E105', 'E106', 'E107', 'E108',  'E109', 'E11', 'E110', 'E111', 'E112', 'E113', 'E114', 'E115', 'E116', 'E117',  'E118', 'E119', 'E12', 'E120', 'E121', 'E122', 'E123', 'E124', 'E125', 'E126',  'E127', 'E128', 'E129', 'E13', 'E130', 'E131', 'E132', 'E133', 'E134', 'E135',  'E136', 'E137', 'E138', 'E139', 'E14', 'E140', 'E141', 'E142', 'E143', 'E144',  'E145', 'E146', 'E147', 'E148', 'E149', 'O240', 'O241', 'O242', 'O243', 'O249' |

Nb. This table displays the ICD codes used for making the baseline tables. Abbreviations; ICD: International classification of disease.

Supplementary table 5: The 5 target phenotypes and the phecodes and the diagnosis codes used to define them

| **Phenotype** | **Phecode** | **ICD codes** |
| --- | --- | --- |
| Heart failure | 428.2 | I50.82, I50.81, I50.8, I50.84. I50.810, I50.812, I50.83, I50.89, I50.814, I50, I50.811, I50.1, I50.813 |
| Aortic valve disease | 394.3 | I106.0, I106.1, I106.2 |
| Mitral valve disease | 394.2 | I05.8, I05.0, I05,I05.2, I05.9, I05.1 |
| Left ventricular hypertrophy | 416 | I51.7 |

### **Supplementary figures**

**Supplementary figure 1:** ROC of the male sex model

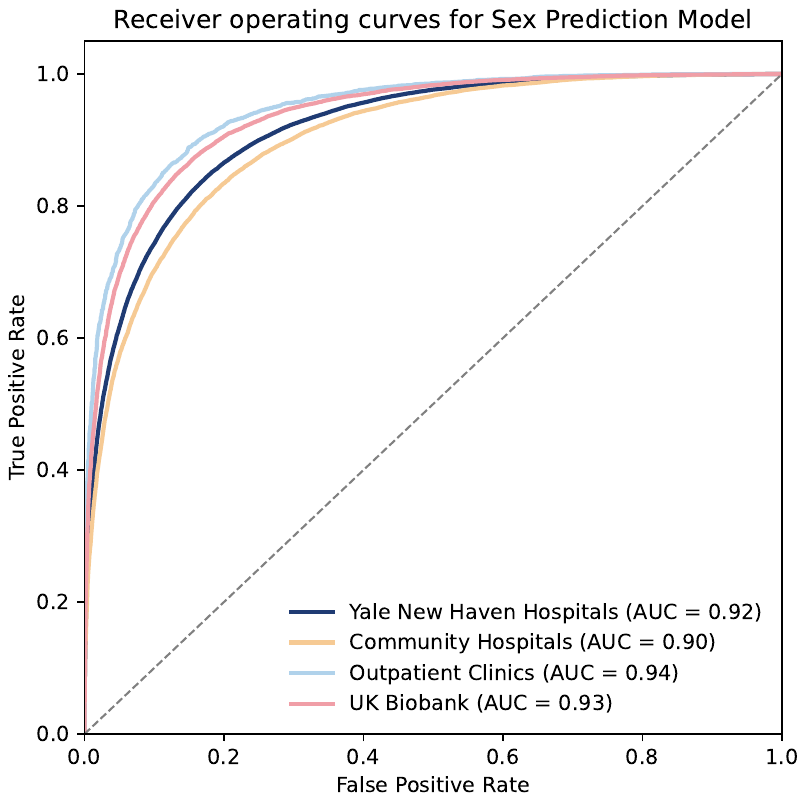

Nb. Receiver Operating Characteristic curves for the sex prediction model across different cohorts: Yale New Haven Hospitals, Community Hospitals, Outpatient Clinics, and UK Biobank. Abbreviations; AUC: Area under the Receiver Operating Curves.

**Supplementary figure 2:** Age and sex distribution of the non-cardiovascular disease phenotypes used to train experimental

models

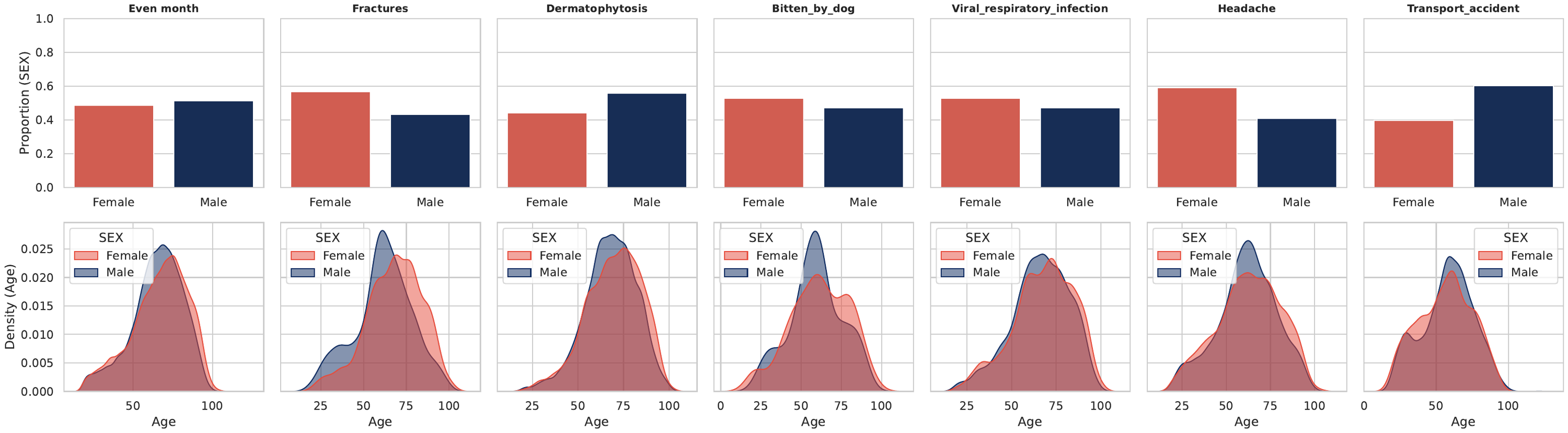

**Supplementary figure 3:** Proportional Hazards and Functional-Form Diagnostics in the YNHH Cohort

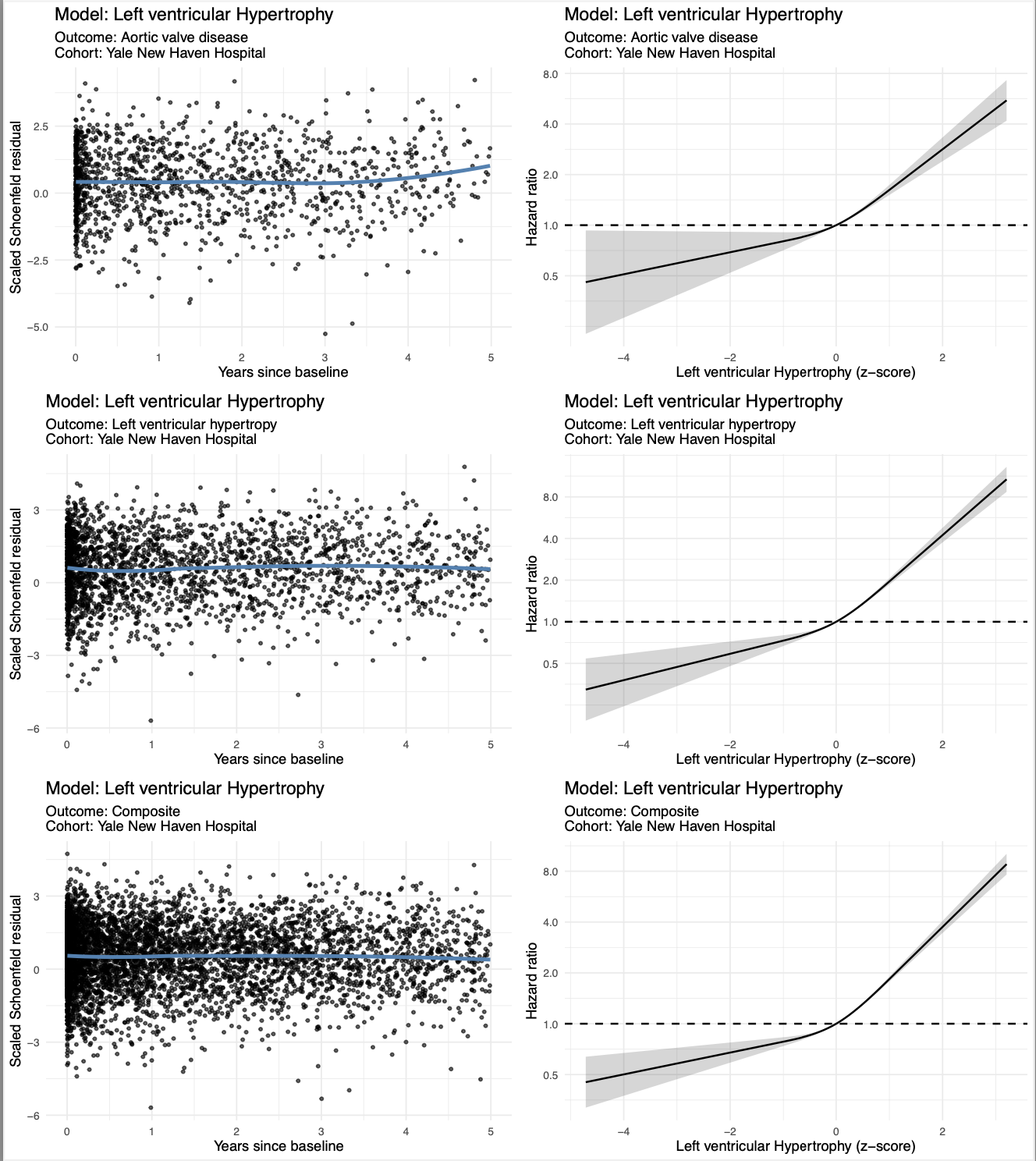

**Supplementary figure 3 *Extension***

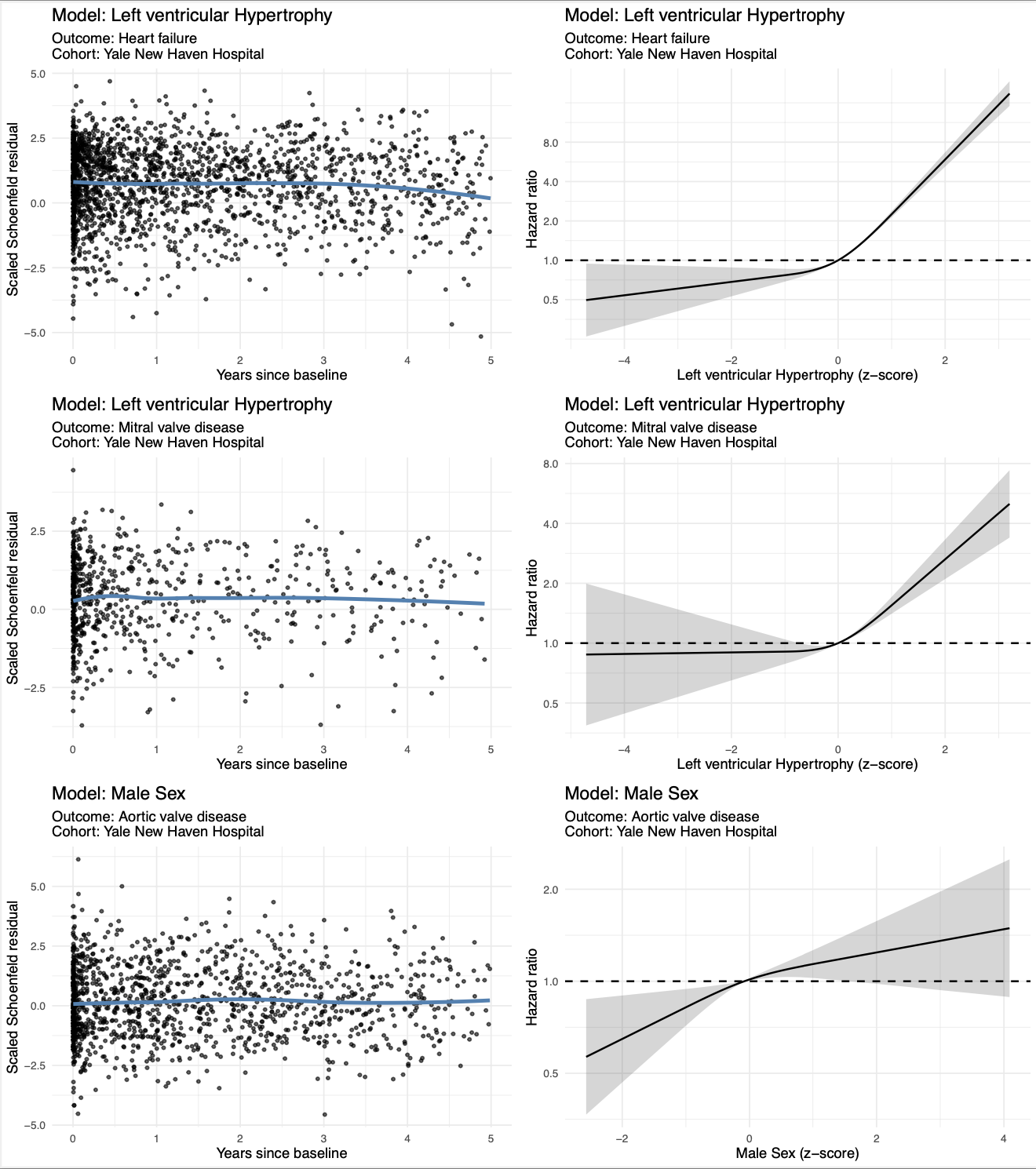

**Supplementary figure 3 *Extension***

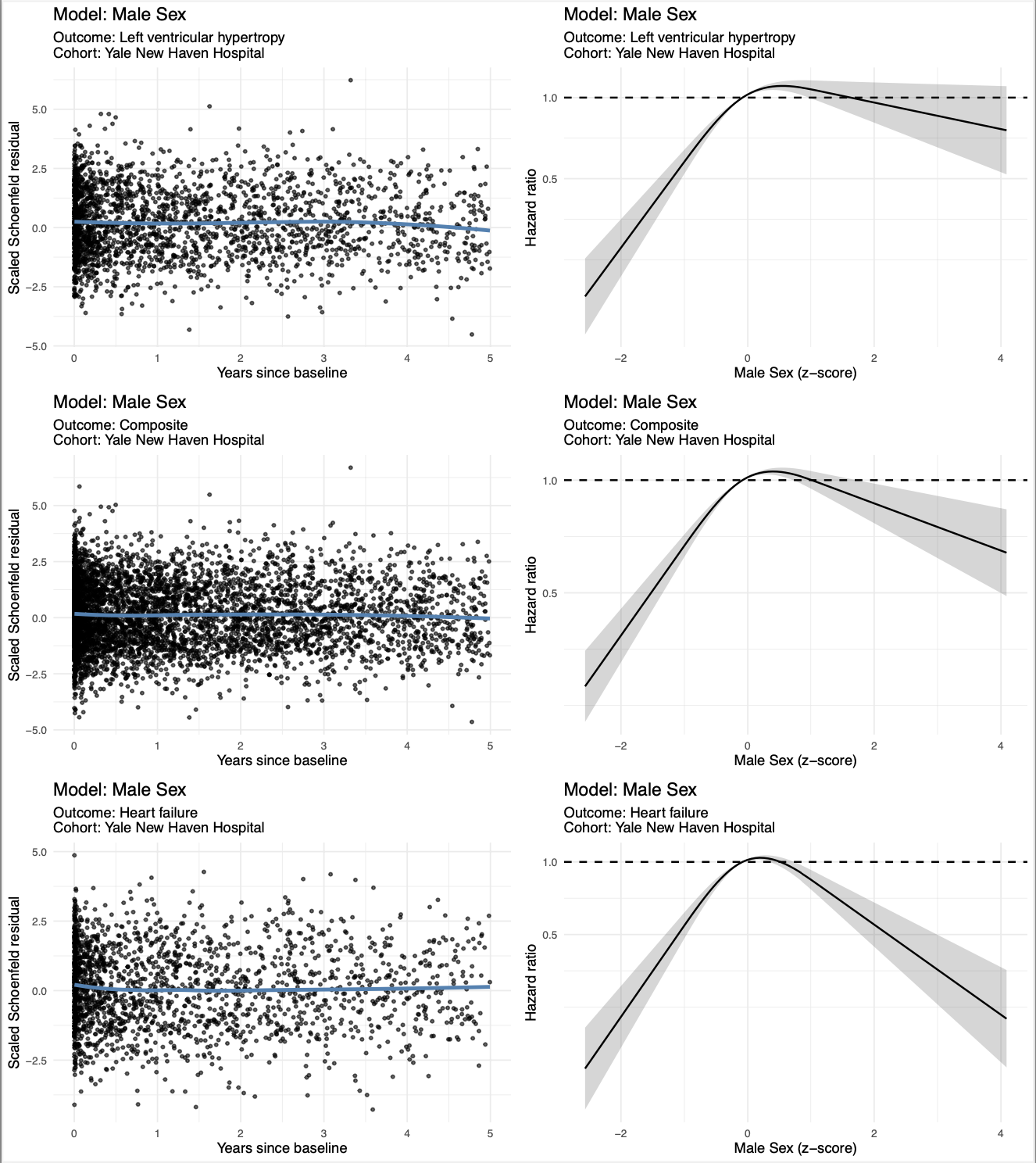

**Supplementary Figure 3 *Extension***
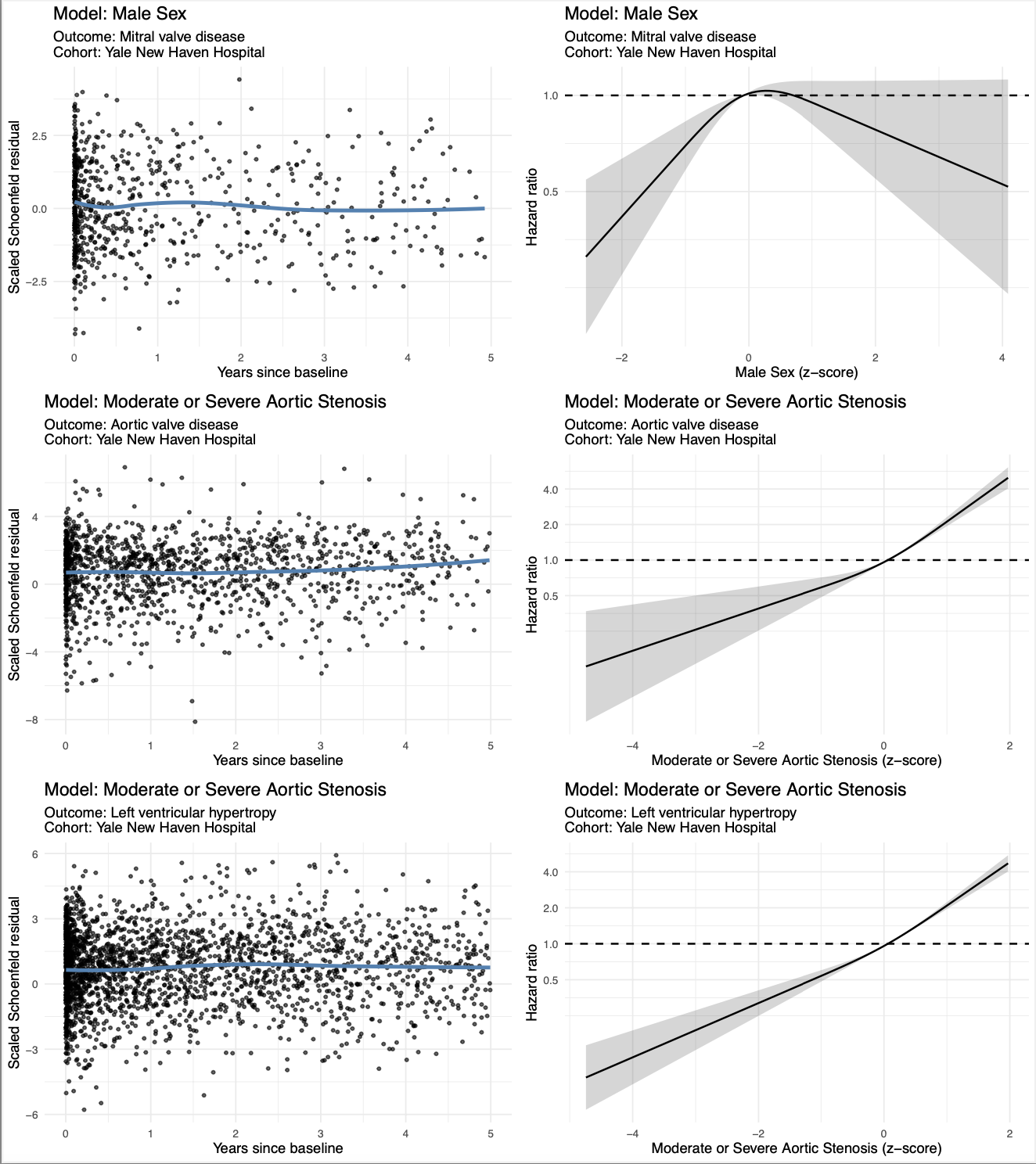

**Supplementary figure 3 *Extension***

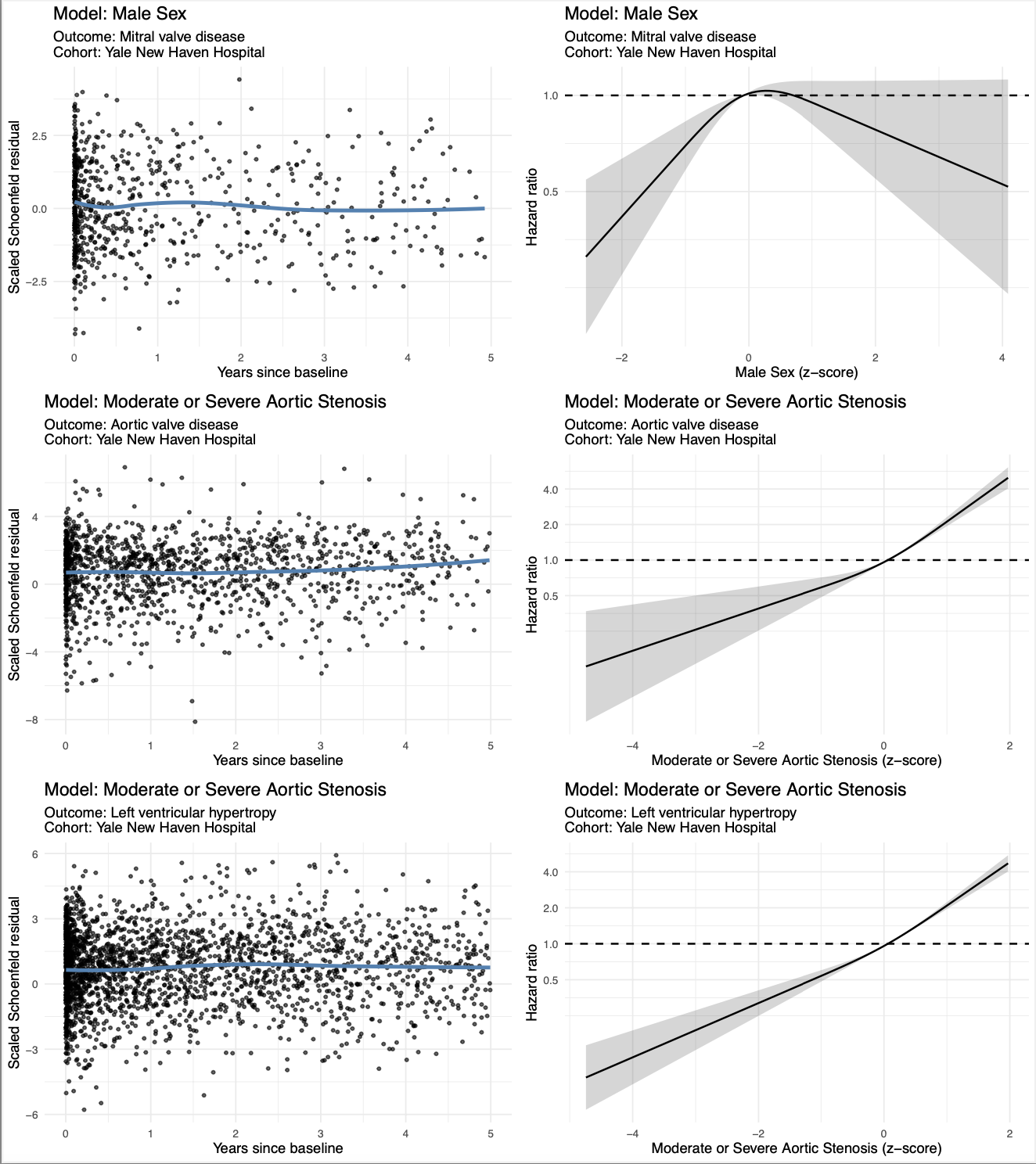

**Supplementary figure 3 *Extension***

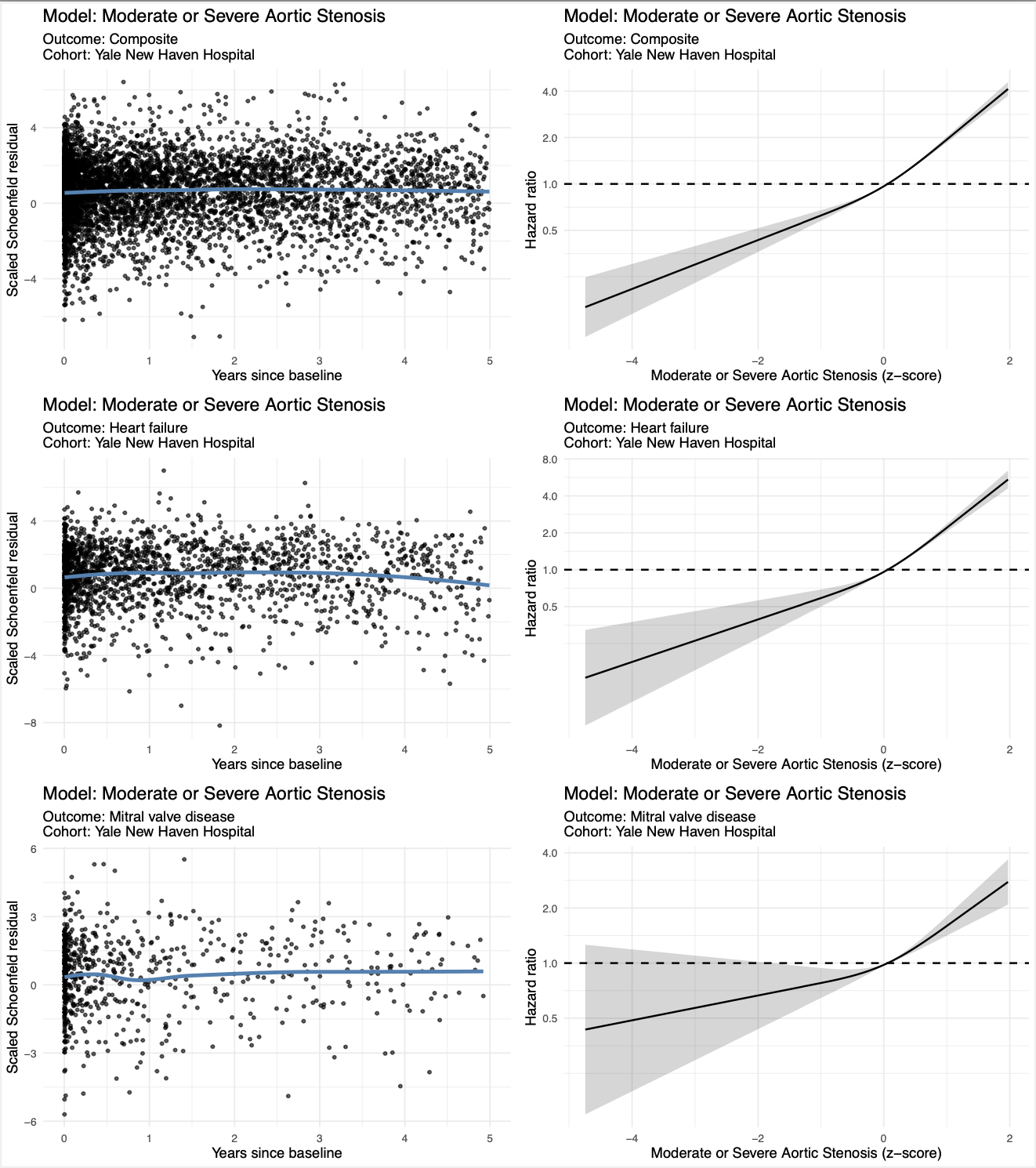

**Supplementary figure 3 *Extension***

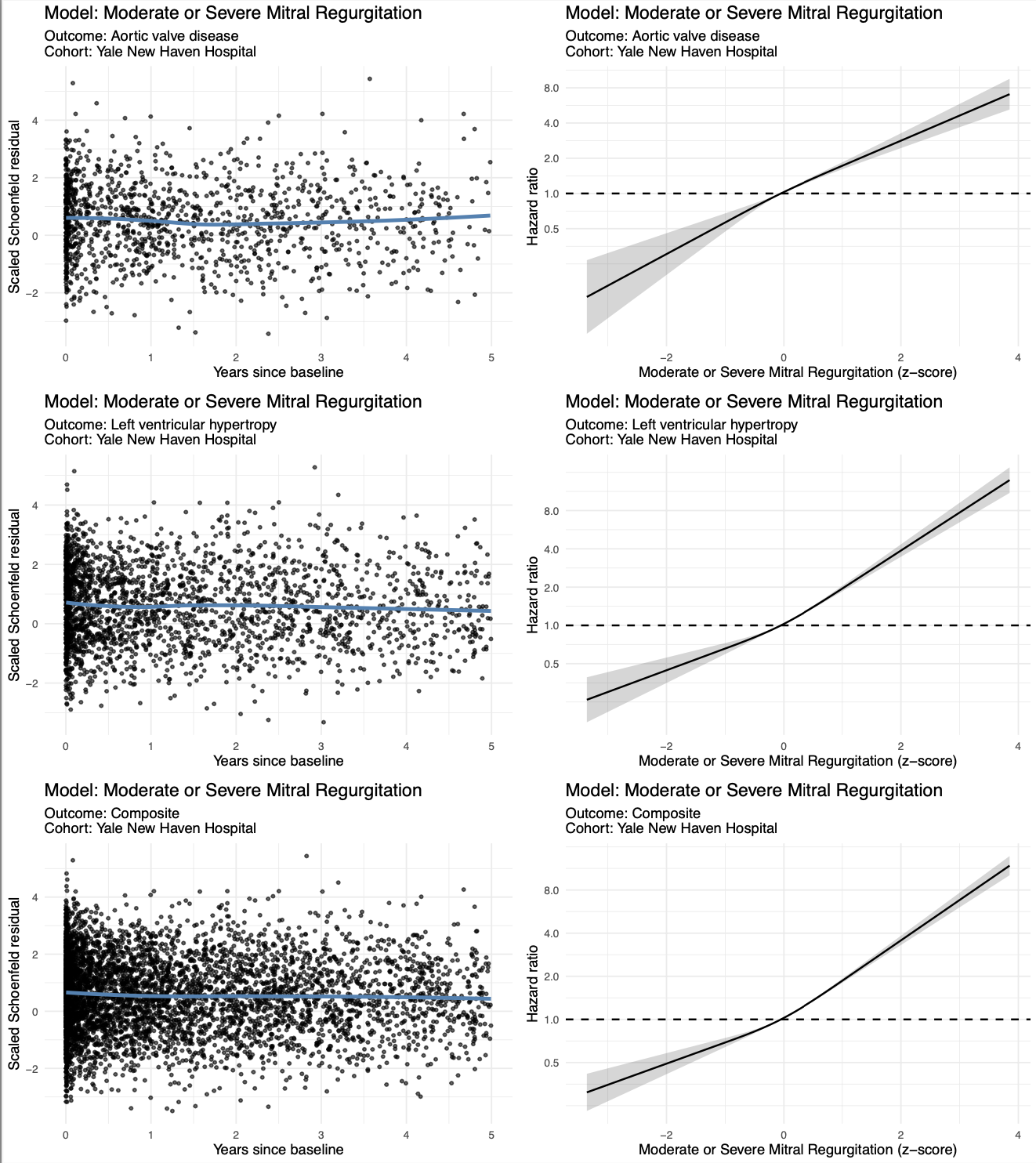

**Supplementary figure 3 *Extension***

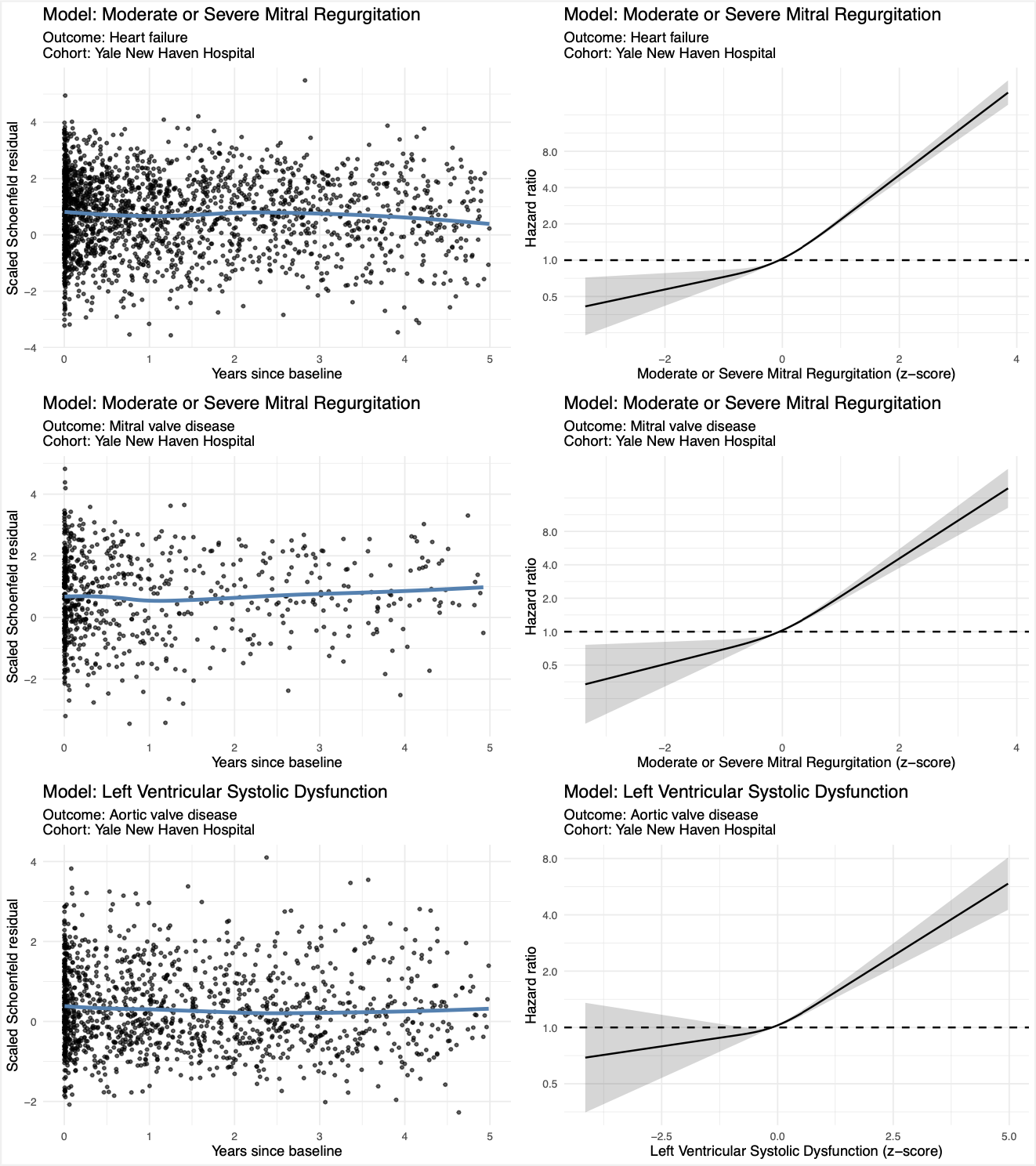

**Supplementary figure 3 *Extension***

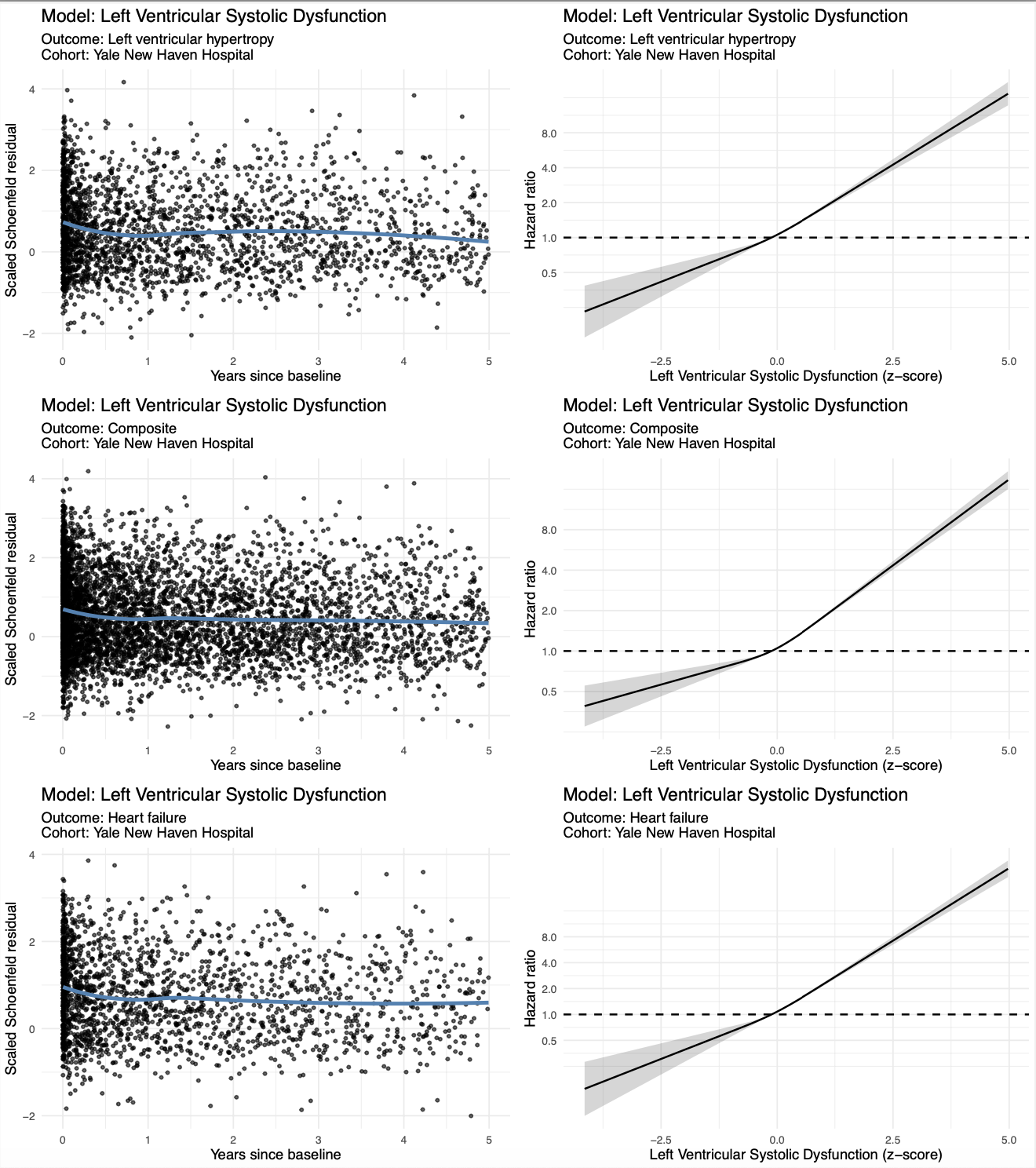

**Supplementary figure 3 *Extension***

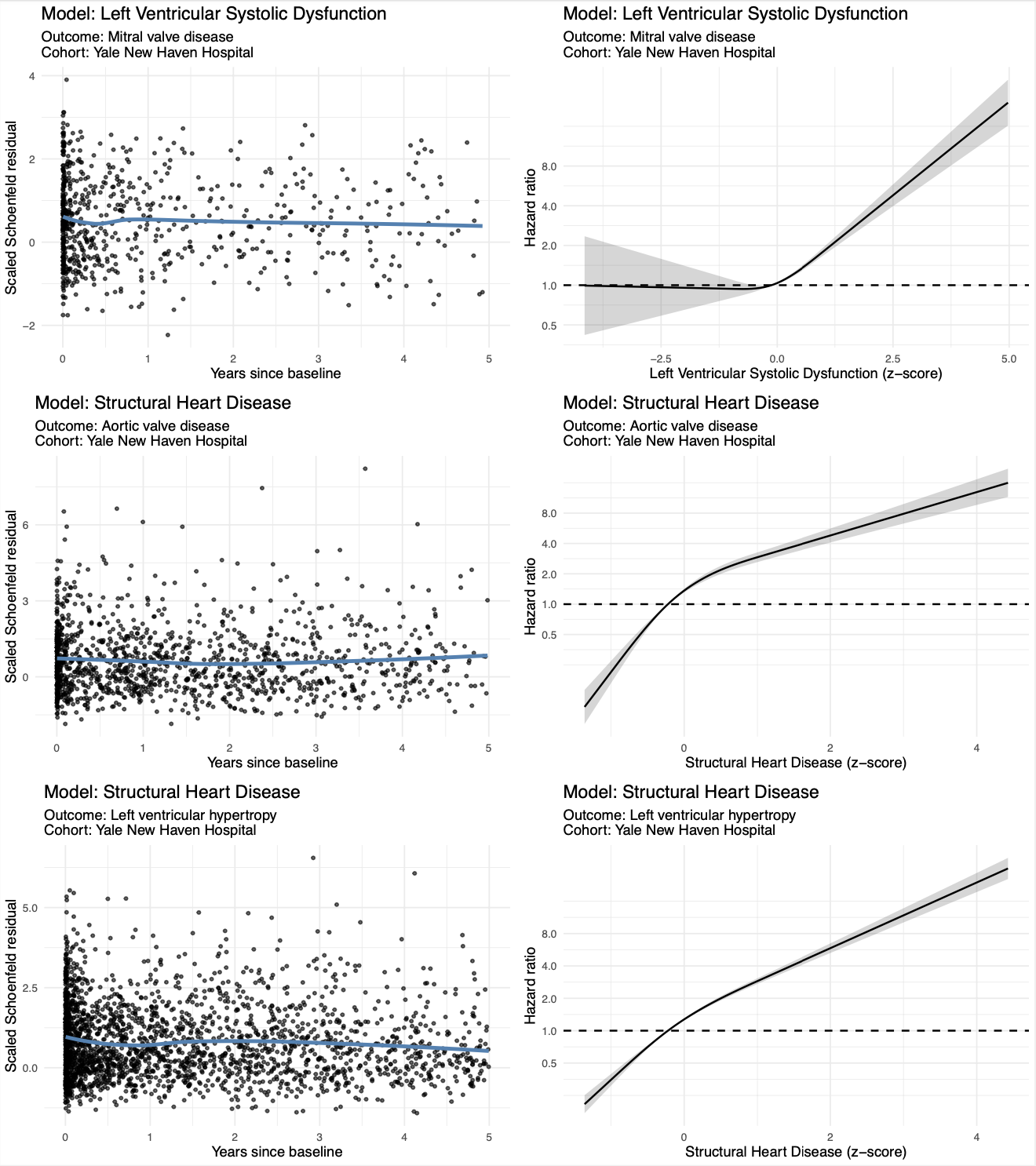

**Supplementary figure 3 *Extension***

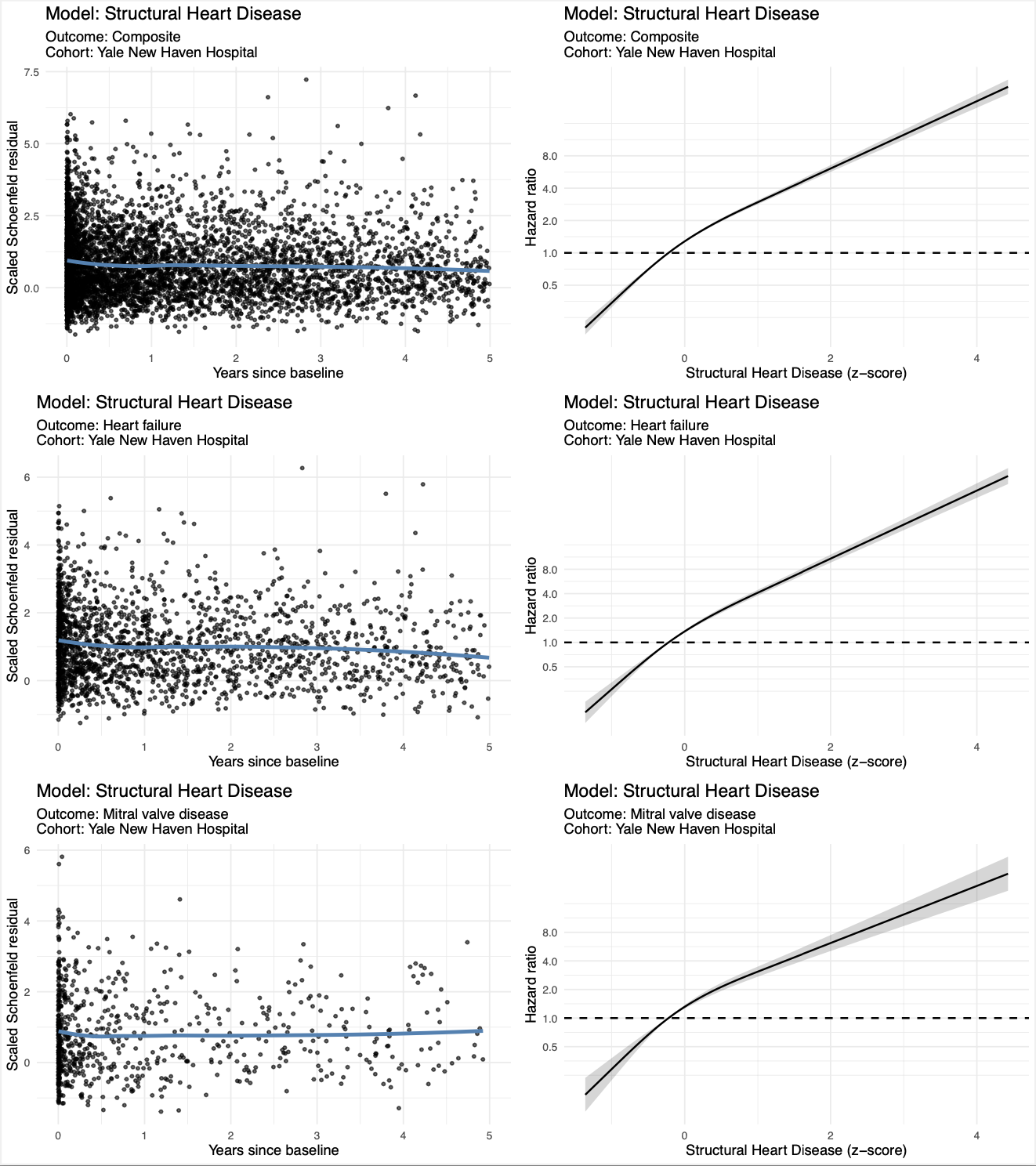

Schoenfeld residuals over time (left) and five-knot restricted cubic spline fits (right) for each AI-ECG predictor, demonstrating no meaningful departures from model assumptions.

**Supplementary figure 4:** Piecharts displaying the distribution of significant phenotypes per group

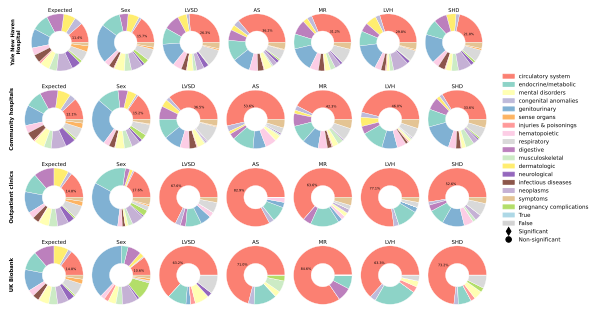

NB. Distribution of significant associations from the phenome-wide association study (PheWAS) for each AI-ECG model across the Yale New Haven Health System, Community Hospitals, Outpatient Clinics, and UK Biobank cohorts. All phenotypes present in the cohort are categorized by group, representing different organ systems or disease categories. Each pie chart represents the proportion of significant phenotype associations with the phenotypes within these groups, as indicated by the color legend. The “Expected” distribution represents the proportion of all tested phenotypes before applying significance thresholds, which represents the distribution as it would be expected by chance. All models but the sex model are more likely to be associated with cardiovascular phenotypes than with any other phenotype, when compared with the expected distribution. Abbreviations: AS, aortic stenosis; HCM, hypertrophic cardiomyopathy; LVH, left ventricular hypertrophy; LVSD, left ventricular systolic dysfunction; MR, mitral regurgitation; SHD, structural heart disease.

**Supplementary figure 5:** Boxplots showing the Odds Ratio’s for enrichment of phenotypes

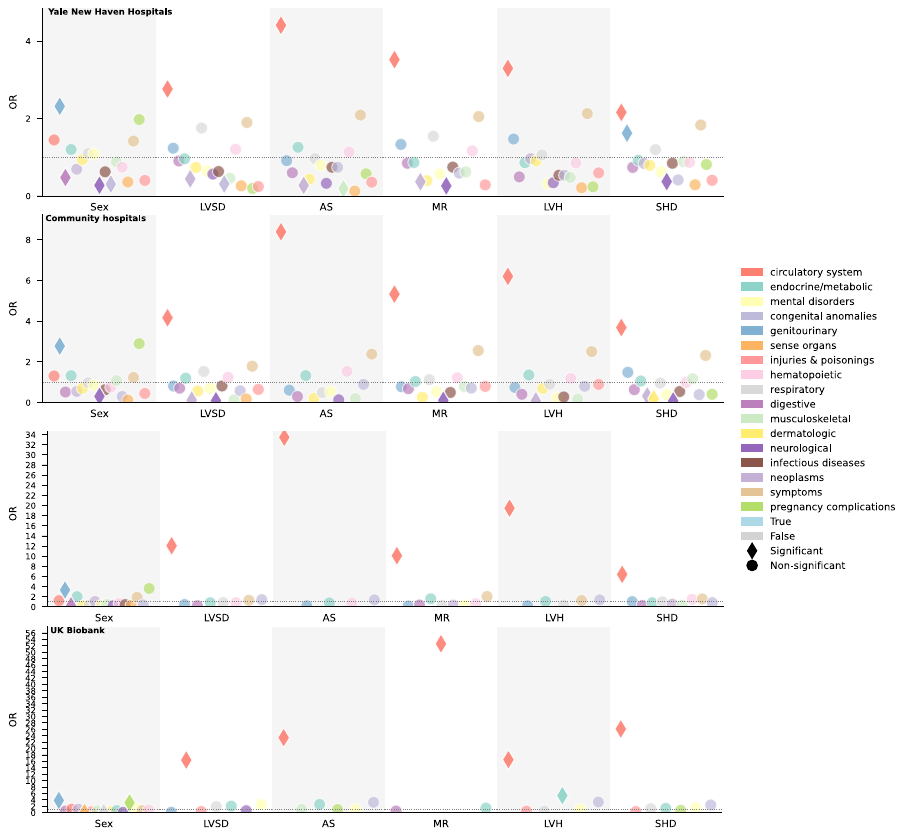

NB. We evaluated whether each disease category was over- or under-represented among the significant PheWAS hits for each AI-ECG model by comparing the observed proportion of significant phenotypes in that category to its proportion among all tested phenotypes. Odds ratios (ORs) were computed from 2×2 contingency tables (observed vs expected) and tested for enrichment using Fisher’s exact test. Each diamond represents the OR for a significant phenotype within a disease category. The y-axis represents the OR, while the x-axis corresponds to different AI-ECG models. Colors correspond to disease categories shown in Panel A as displayed in the legend. Abbreviations: AS, aortic stenosis; LVH, left ventricular hypertrophy; LVSD, left ventricular systolic dysfunction; MR, mitral regurgitation; SHD, structural heart disease; OR: odds ratio.

**Supplementary figure 6:** Volcano plots of the associations of all the models in all four cohorts

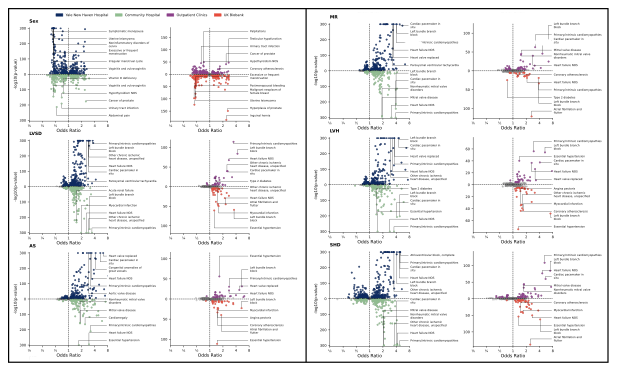

Nb. Volcano plots displaying the associations between various phenotypes and AI-ECG model predictions across four cohorts: Yale New Haven Hospital (YNHH, blue), Community Hospitals (green), Outpatient Clinics (red), and UK Biobank (purple). Each panel represents a different AI-ECG model prediction, including Sex, Mitral Regurgitation (MR), Left Ventricular Systolic Dysfunction (LVSD), Left Ventricular Hypertrophy (LVH), Aortic Stenosis (AS), and Structural Heart Disease (SHD). The x-axis represents the odds ratio , and the y-axis shows the -log₁₀(p-value). Labeled points indicate the top 6 statistically significant associations. Abbreviations; AS: Aortic Stenosis, LVH: Left Ventricular Hypertrophy, LVSD: Left Ventricular Systolic Dysfunction, MR: Mitral Regurgitation, SHD: Structural Heart Disease.

**Supplementary figure 7:** Sensitivity analysis to assess the effect of concomitant diseases on the overlap in phenotypic associations
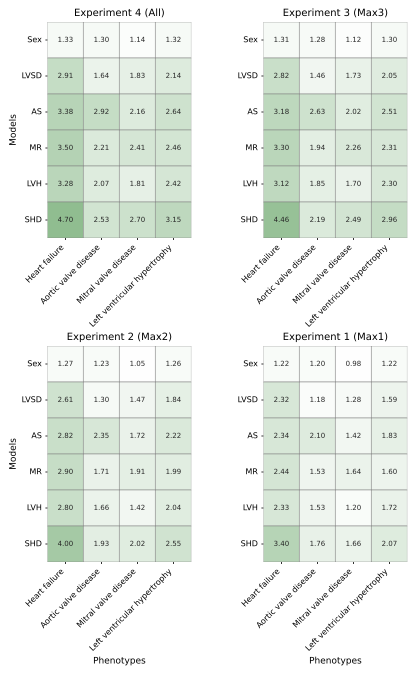

Odds ratios for AI-ECG model predictions and key phenotypes across four experiments. In Experiment 1, individuals with more than one of the four target phenotypes were excluded; in Experiment 2, those with more than two; in Experiment 3, those with more than three. Experiment 4 replicates the main analysis without filtering for overlapping target phenotypes. While the effect sizes are slightly modestly attenuated in a more restricted population, the non-specificity selectivity of the phenotypic associations remains. Abbreviations: AS: Aortic stenosis, LVH: Left ventricular hypertrophy, LVSD: Left ventricular systolic dysfunction, MR: Mitral regurgitation, SHD: Structural heart disease composite model, Sex: Biological sex prediction model.

**Supplementary Figure 8:** Exploration of the phenotypic associations of hypertrophic subphenotypes

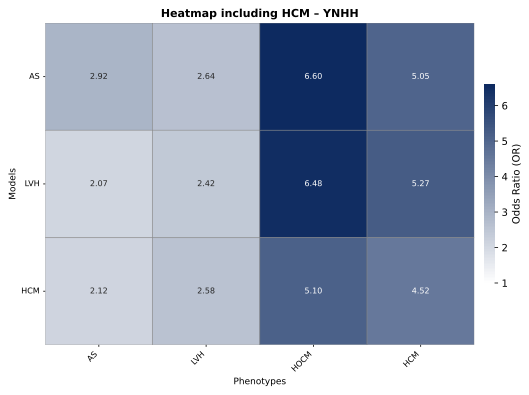

This heatmap displays odds ratios from age- and sex-adjusted logistic regression analyses between AI-ECG models trained to detect aortic stenosis, hypertrophic cardiomyopathy, and hypertrophic obstructive cardiomyopathy, and hypertrophic phenotypes. Despite being trained on distinct labels, the models show substantial overlap in their phenotypic associations. Abbreviations: AS: aortic stenosis, LVH: left ventricular hypertrophy, HCM: hypertrophic cardiomyopathy, HOCM: Hypertrophic obstructive cardiomyopathy.

**Supplementary Figure 9:** Scatterplot of Odds Ratio’s comparing Yale New Haven Health to the community hospitals

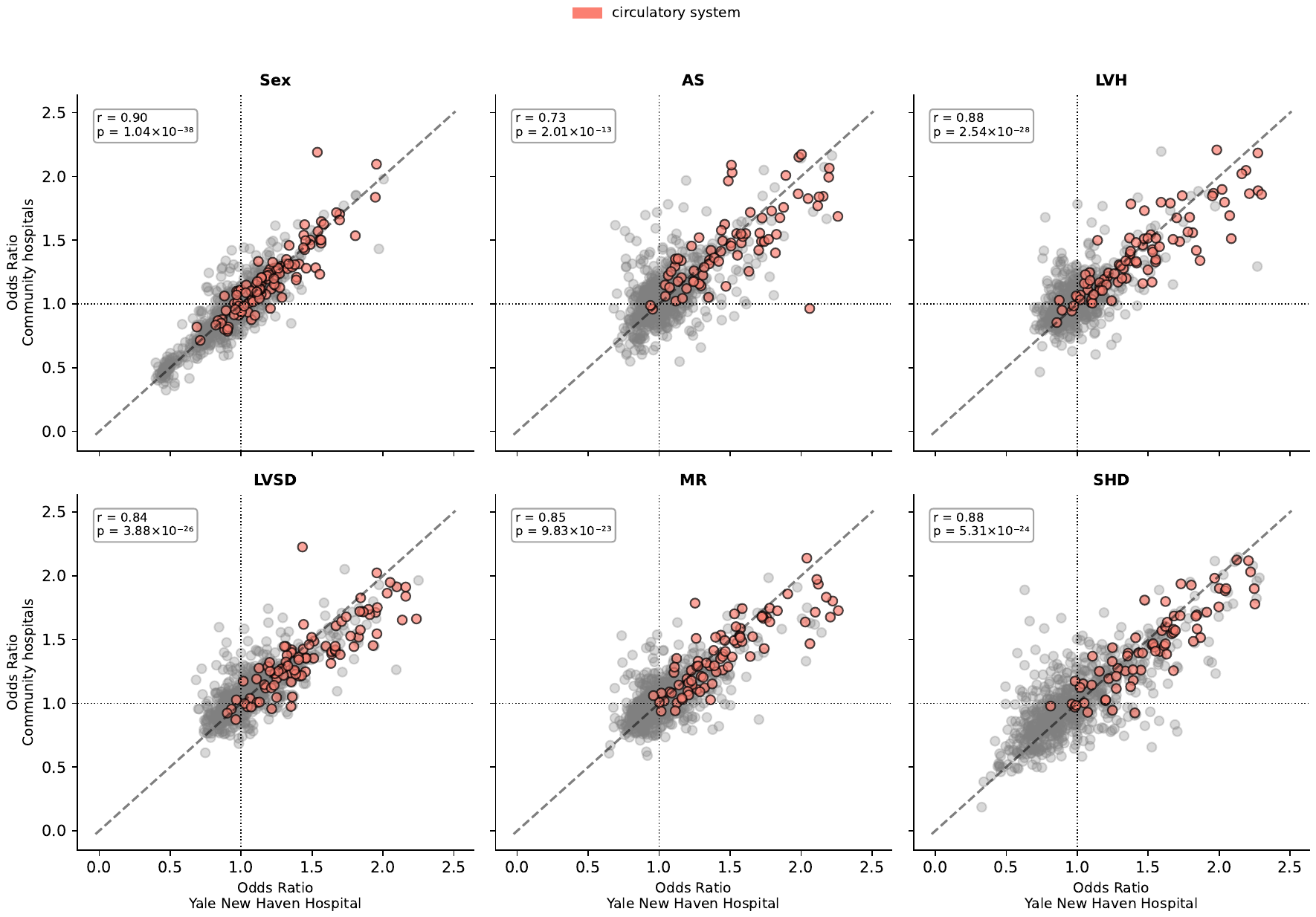

Nb. This figure presents scatter plots comparing odds ratios (ORs) derived from Yale New Haven Hospital (YNHH) with those from the four community hospitals. Each subplot represents a different AI-ECG model predicting a cardiovascular phenotype, including aortic stenosis, left ventricular hypertrophy, left ventricular systolic dysfunction, mitral regurgitation, structural heart disease, and male sex prediction. The diagonal dashed line represents the identity line, indicating perfect agreement between YNHH and the compared cohort. Abbreviations; AS: Aortic Stenosis, LVH: Left Ventricular Hypertrophy, LVSD: Left Ventricular Systolic Dysfunction, MR: Mitral Regurgitation, SHD: Structural Heart Disease.

**Supplementary Figure 10:** Scatterplot of Odds Ratio’s comparing Yale New Haven Health to the outpatient clinics

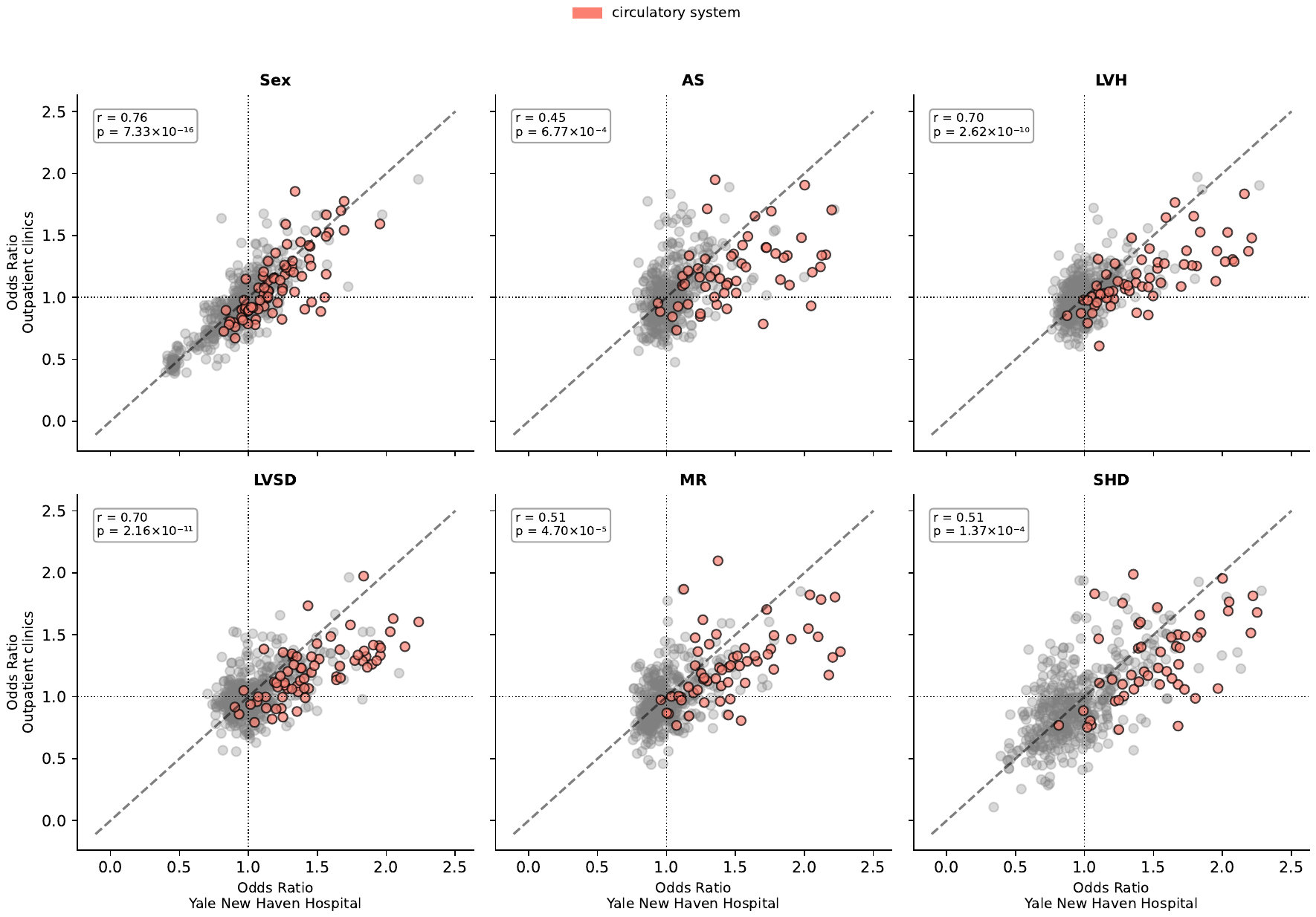

Nb. This figure presents scatter plots comparing odds ratios (ORs) derived from Yale New Haven Hospital (YNHH) with those from the outpatient clinic hospitals. Each subplot represents a different AI-ECG model predicting a cardiovascular phenotype, including aortic stenosis, left ventricular hypertrophy, left ventricular systolic dysfunction, mitral regurgitation, structural heart disease, and male sex prediction. The diagonal dashed line represents the identity line, indicating perfect agreement between YNHH and the compared cohort. Abbreviations; AS: Aortic Stenosis, LVH: Left Ventricular Hypertrophy, LVSD: Left Ventricular Systolic Dysfunction, MR: Mitral Regurgitation, SHD: Structural Heart Disease.

**Supplementary Figure 11:** Scatterplot of Odds Ratio’s comparing Yale New Haven Health to the UK Biobank

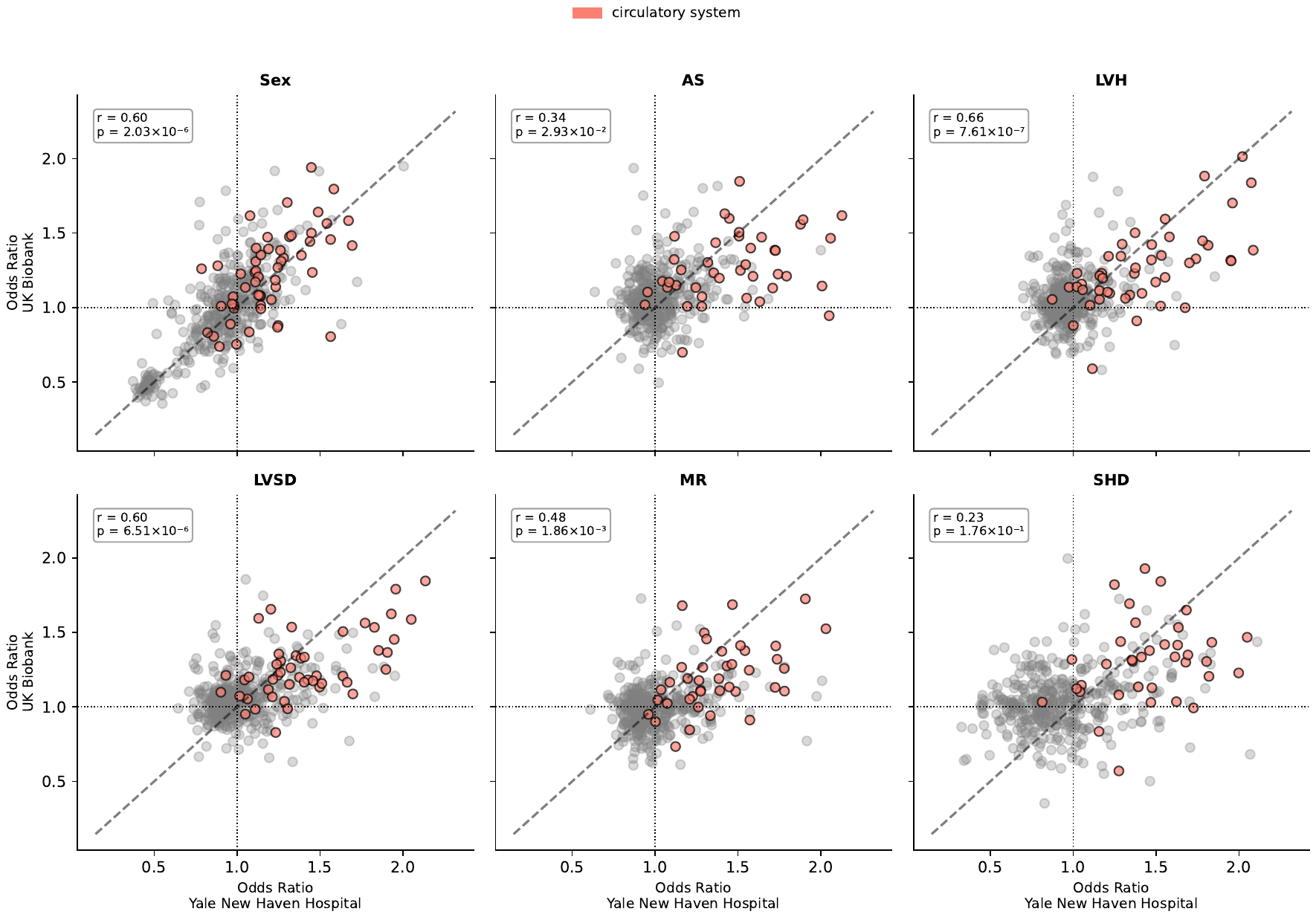

Nb. This figure presents scatter plots comparing odds ratios (ORs) derived from Yale New Haven Hospital (YNHH) with those from three different clinical settings: community hospitals, outpatient clinics, and UK Biobank. Each subplot represents a different AI-ECG model predicting a cardiovascular phenotype, including aortic stenosis, left ventricular hypertrophy, left ventricular systolic dysfunction, mitral regurgitation, structural heart disease, and male sex prediction. The diagonal dashed line represents the identity line, indicating perfect agreement between YNHH and the compared cohort. Abbreviations; AS: Aortic Stenosis, LVH: Left Ventricular Hypertrophy, LVSD: Left Ventricular Systolic Dysfunction, MR: Mitral Regurgitation, SHD: Structural Heart Disease.

**Supplementary figure 12:** Odds ratios of AI-ECG model probabilities with on-target phenotype profiles across AI-ECG models in four independent cohorts

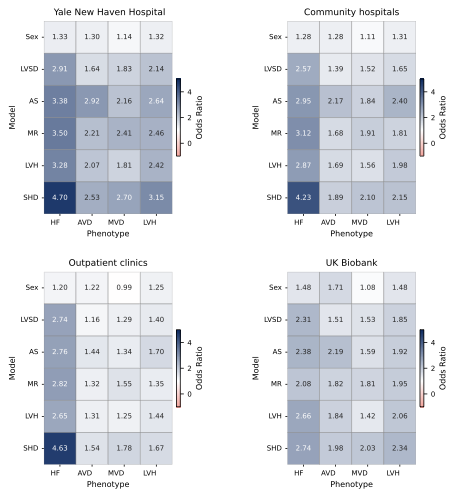

This heatmap displays odds ratios derived from the phenome wide association study, between AI-ECG outputs and the target cardiovascular phenotypes in all four included cohorts. Across models, the strongest phenotypic association was with heart failure. Of all models, the SHD model exhibited stronger associations compared to the single disease models. Phenotypic patterns are highly similar between models, a pattern that is consistent across cohorts. Abbreviations: HF, Heart Failure; AVD, Aortic Valve Disease; MVD, Mitral Valve Disease; TVD, Tricuspid Valve Disease; LVH, Left Ventricular Hypertrophy; AS, Aortic Stenosis model; LVSD, Left Ventricular Systolic Dysfunction model; MR, Mitral Regurgitation model; SHD, Structural Heart Disease model; Sex, Biological Sex prediction model, OR: Odds ratio.

**Supplementary figure 13:** Correlation of on-target phenotype association profiles across AI-ECG models in four independent cohorts

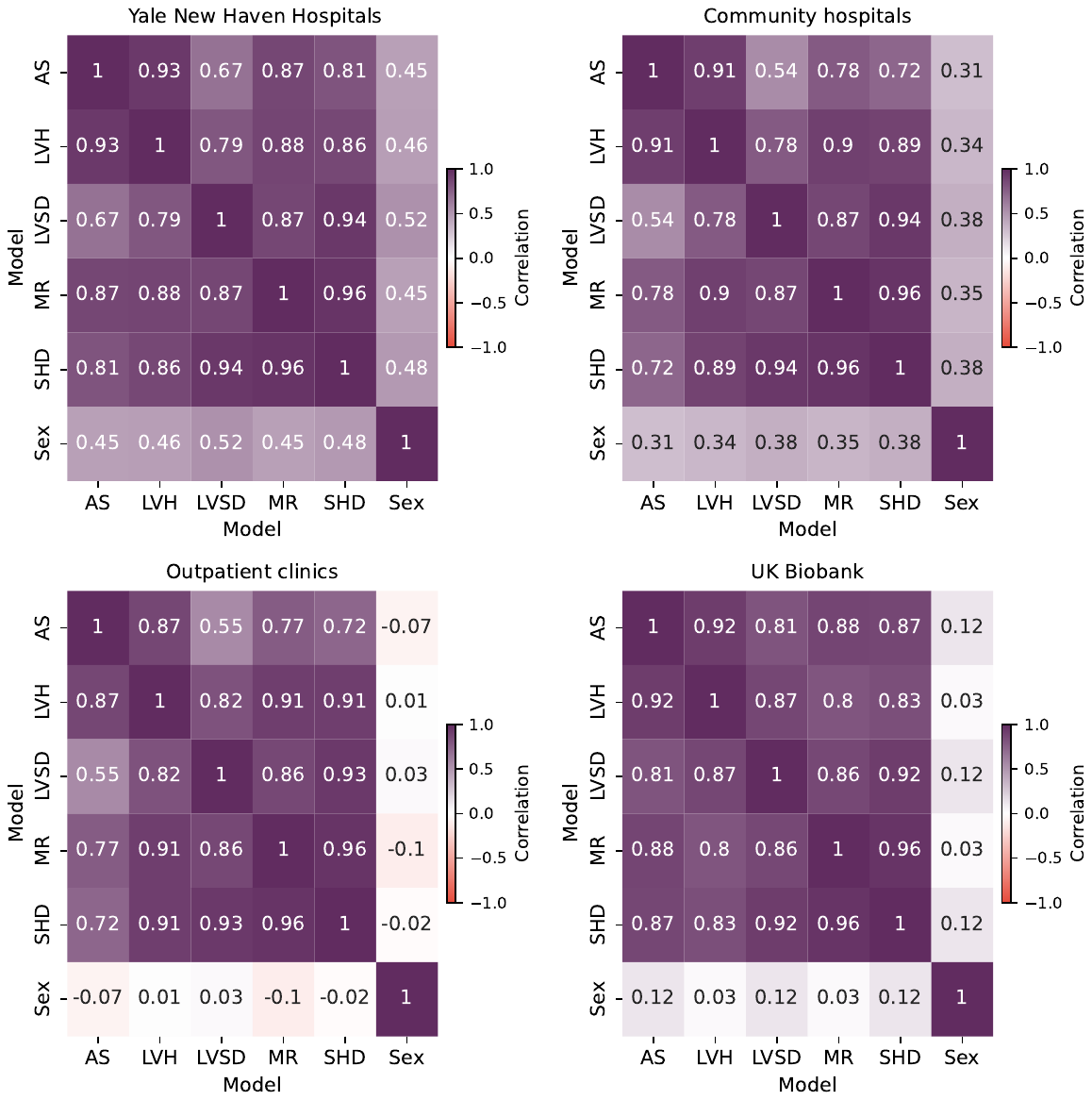

N.B. These heatmaps show Pearson correlation coefficients between odds ratio (OR) profiles across all phenotypes in the cardiovascular group. Correlations are shown for four cohorts: Yale New Haven Hospitals, community hospitals, outpatient clinics, and the UK Biobank. Strong correlations were observed among cardiovascular disease (CVD) models, with the SHD model consistently showing high similarity with other disease-specific models. The model trained to predict biological sex showed consistently weaker correlation with cardiovascular models, particularly in the UK Biobank where inverse correlations were observed. AS: Aortic stenosis, LVH: Left ventricular hypertrophy, LVSD: Left ventricular systolic dysfunction, MR: Mitral regurgitation, SHD: Structural heart disease composite model, Sex: Biological sex prediction model.

**Supplementary Figure 14:** Time to event analysis for the Sex and LVSD model

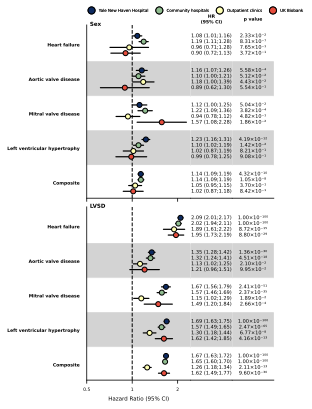

NB. This forest plot displays hazard ratios (HRs) and 95% confidence intervals (CIs) for the Sex and Left Ventricular Systolic Dysfunction (LVSD) AI-ECG models across multiple cardiovascular outcomes, derived from age- and sex-adjusted Cox proportional hazards models. Results are shown separately for each healthcare setting: Yale New Haven Hospitals, community hospitals, outpatient clinics, and the UK Biobank (UKB). Abbreviations: HR: Hazard ratio, CI: Confidence interval, LVSD: Left ventricular systolic dysfunction, UKB: UK Biobank.

**Supplementary Figure 15:** Time to event analysis for the AS and MR model

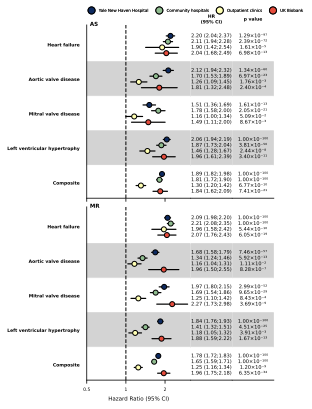

NB. This forest plot displays hazard ratios (HRs) and 95% confidence intervals (CIs) for the Aortic stenosis (AS) and Mitral regurgitation (MR) AI-ECG models across multiple cardiovascular outcomes, derived from age- and sex-adjusted Cox proportional hazards models. Results are shown separately for each healthcare setting: Yale New Haven Hospitals, community hospitals, outpatient clinics, and the UK Biobank (UKB). Abbreviations: HR: Hazard ratio, CI: Confidence interval, AS: aortic stenosis, MR: mitral regurgitation, UKB: UK Biobank.

**Supplementary Figure 16:** Time to event analysis for the LVH and SHD model

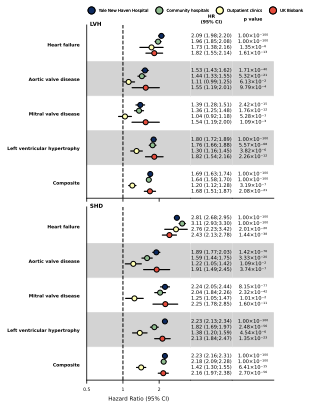

NB. This forest plot displays hazard ratios (HRs) and 95% confidence intervals (CIs) for the left ventricular hypertrophy (LVH) and structural heart disease (SHD) model AI-ECG models across multiple cardiovascular outcomes, derived from age- and sex-adjusted Cox proportional hazards models. Results are shown separately for each healthcare setting: Yale New Haven Hospitals, community hospitals, outpatient clinics, and the UK Biobank (UKB). Abbreviations: HR: Hazard ratio, CI: Confidence interval, LVH: left ventricular hypertrophy, SHD: structural heart disease, UKB: UK Biobank.

**Supplementary figure 17:** Associations between experimental model outputs and cross-sectional disease phenotypes

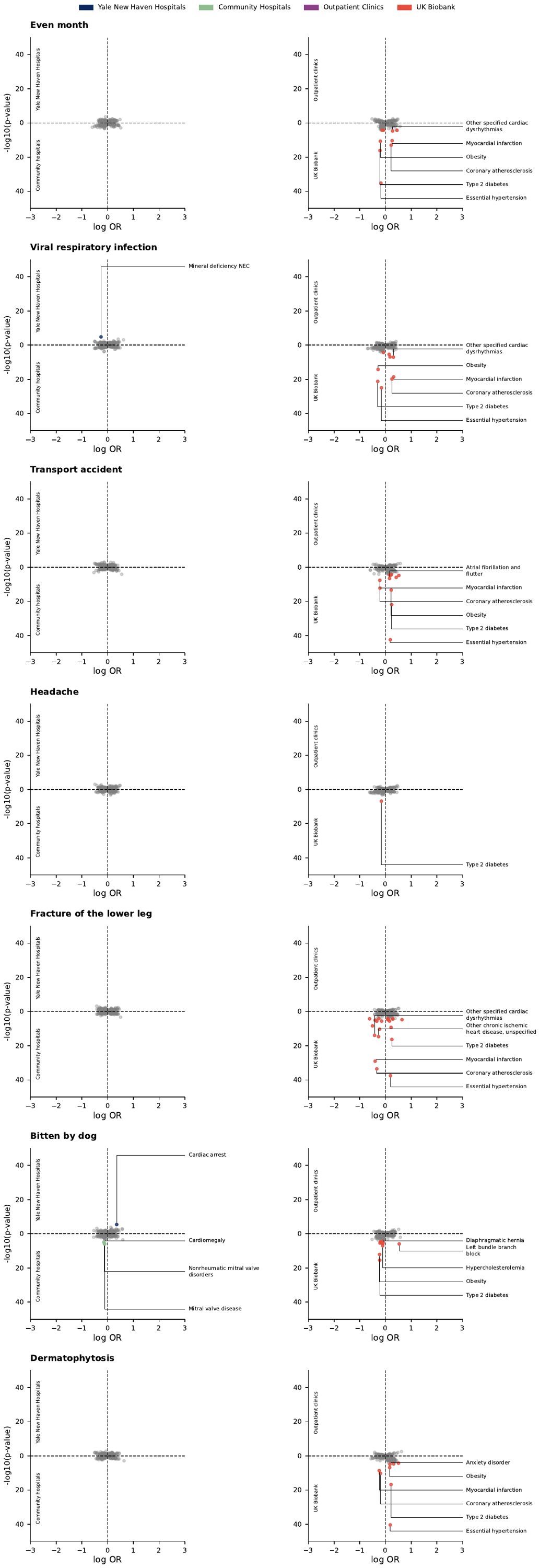

NB This plot displays the odds ratio’s and -log10(p) values that resulted from logistic regression between the model outputs of experimental models and cross-sectional phenotypes. Abbreviations: OR: Odds ratio.

**Supplementary Figure 18:** Associations between experimental model outputs and cross-sectional disease phenotypes

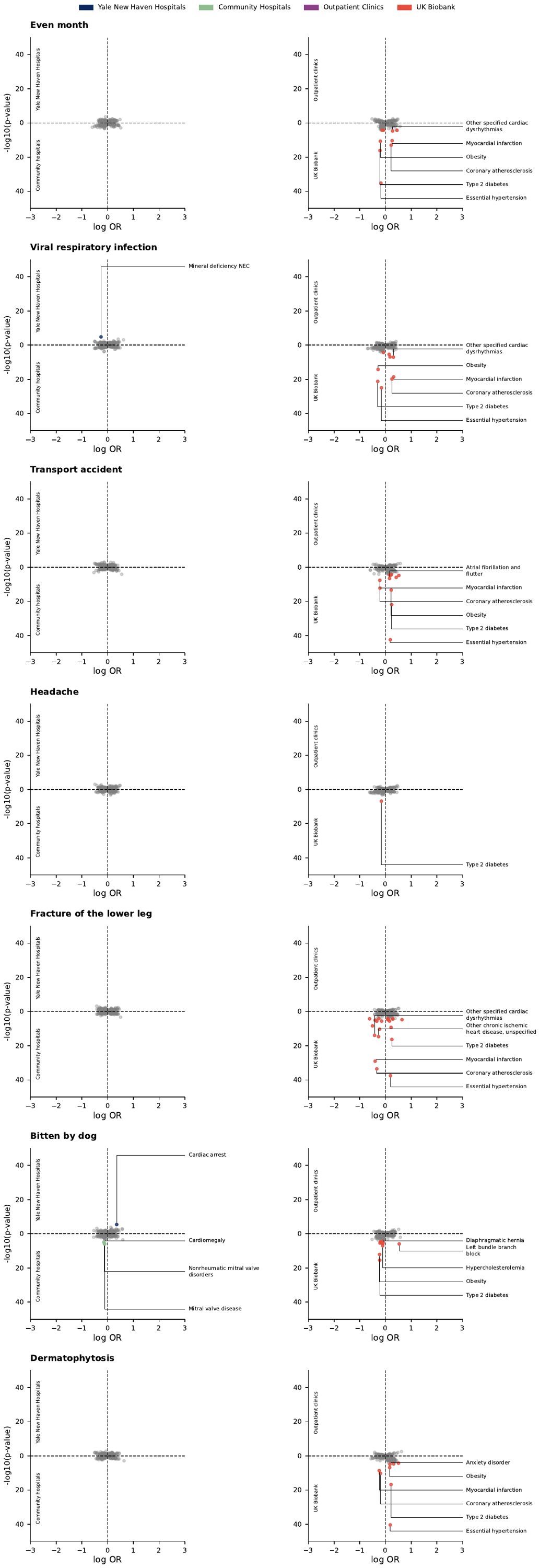

NB This plot displays the odds ratio’s and -log10(p) values that resulted from logistic regression between the model outputs of experimental models and cross-sectional phenotypes. Abbreviations: OR: Odds ratio.
